# Mental health of patients with mental illness during the COVID-19 pandemic lockdown: A questionnaire-based survey weighted for attrition

**DOI:** 10.1101/2021.03.13.21253363

**Authors:** Pernille Kølbæk, Oskar Hougaard Jefsen, Maria Speed, Søren Dinesen Østergaard

**Author notes:** Corresponding author: Pernille Kølbæk, Department of Affective Disorders, Aarhus University Hospital—Psychiatry, Palle Juul-Jensens Boulevard 175, DK–8200 Aarhus N, Denmark.

## Abstract

**Importance:** Individuals with pre-existing mental illness may be particularly vulnerable to the negative impact that the coronavirus disease 2019 (COVID-19) pandemic seems to have on mental health. Most prior studies on this topic are limited by non-random sampling, lacking information on non-respondents, and self-reporting of mental illness. In the present study, we aimed to overcome these limitations via random sampling, acquisition of clinical and sociodemographic data on both respondents and non-respondents, and weighting of results informed by attrition.

**Objective:** To assess whether patients with mental illness experienced deterioration in mental health during the nationwide COVID-19 lockdown of Denmark in the Spring of 2020.

**Design:** A cross-sectional, questionnaire-based survey coupled with sociodemographic and clinical data from the medical records of all invitees. The latter enabled analysis of attrition and weighting of results.

**Setting:** The psychiatric services of the Central Denmark Region.

**Participants:** A total of 992 randomly drawn patients diagnosed with mental illness in the psychiatric services of the Central Denmark Region prior to the lockdown responded to the online survey (response rate of 21.6%).

**Exposure:** The four-week nationwide lockdown during the COVID-19 pandemic (from March 11 to April 15, 2020).

**Main Outcomes and Measures:** The online questionnaire included the 18-item Brief Symptom Inventory (BSI-18), the five-item World Health Organization Well-Being Index (WHO-5), and 14 questions evaluating worsening or improvement in symptoms during the lockdown using the pre-pandemic period as reference. Perceived reasons for deterioration of mental health were also reported.

**Results:** The weighted mean WHO-5 and BSI-18 scores were 38 and 28, respectively. A total of 52% of the respondents reported that their mental health had deteriorated during the lockdown, while 33% reported no change, and 16% reported improvement. The most commonly reported reasons for deterioration were loneliness, disruption of routines, concerns about coronavirus, less frequent contact with family/friends, boredom, and reduced access to psychiatric care.

**Conclusion and Relevance:** More than half of the patients with mental illness reported worsening of their mental health during the pandemic lockdown. There should be increased emphasis on ensuring both social and clinical support for individuals with mental illness during pandemics.

**KEY POINTS:** *Questions:* Did patients with mental illness experience worsening of their mental health during the nationwide COVID-19 lockdown of Denmark in the Spring of 2020?

*Findings:* Of the 925 respondents, 52% reported that their mental health had deteriorated during the lockdown, while 33% reported no change and 16% reported improvement. The most commonly reported reasons for deterioration were loneliness and disruption of routines.

*Meaning:* These findings suggest that there should be increased emphasis on ensuring both social and clinical support for individuals with mental illness during the ongoing and potential future pandemics.

## INTRODUCTION

The coronavirus disease 2019 (COVID-19) pandemic is taking a tremendous toll on societies across the globe. The risk of contracting COVID-19, the experience of having COVID-19, and societal restrictions to reduce the spread of the virus, including lockdowns, social distancing, and preventive isolation, may all have detrimental effects on mental health.^1,2^ Several studies have investigated the mental health effects of the COVID-19 pandemic at the general population level, with the vast majority reporting that the pandemic seems to have had a negative impact.^3-7^

Individuals with mental illness may be particularly vulnerable to the impact of the COVID-19 pandemic. However, the relatively few studies exploring whether individuals with mental illness have experienced exacerbation during the pandemic have reached mixed results. While some have reported that the pandemic has caused deterioration of mental health,^8-16^ others suggest a neutral effect^6,17-19^ or even improvement in mental health among specific subgroups of patients.^14,20,21^ This literature is, however, marred by several methodological limitations, including the use of non-random sampling,^10,11,16-20^ self-reported diagnoses,^10,11,16,21^ and a lack of information on those choosing not to participate,^6,8,12,13,15-21^ which likely results in selection bias, reporting bias, and suboptimal generalizability of the findings.^22^ We therefore designed a questionnaire-based survey aimed at overcoming these limitations via random sampling in a population of patients diagnosed in psychiatric services for whom clinical and sociodemographic information from electronic health records was available for both respondents and non-respondents. The latter enabled attrition analysis and weighting of results. The study addressed the following research questions:

I. How did the nationwide COVID-19 lockdown of Denmark in the Spring of 2020 affect the mental health of patients with mental illness?
II. Which clinical and sociodemographic characteristics were associated with deterioration of mental health among patients with mental illness during the nationwide COVID-19 lockdown of Denmark in the Spring of 2020?

## METHODS

### Design

We conducted a cross-sectional, questionnaire-based, online survey coupled with sociodemographic and clinical data from the electronic medical records of all invitees, which enabled attrition analysis and weighting of results. The description of the methods and results align with the reporting guidelines of the American Association for Public Opinion Research.^23^

### Setting

The study was conducted in the psychiatric services of the Central Denmark Region. The Central Denmark Region is the second largest of the five Danish Regions with a catchment area of approximately 1.3 million people. The psychiatric services of the Central Denmark Region comprise five publicly funded psychiatric hospitals providing cost-free inpatient, outpatient, and emergency psychiatric treatment.

### The nationwide COVID-19 lockdown of Denmark in the Spring of 2020

The first case of COVID-19 in Denmark was reported on February 26, 2020.^24^ On March 11, 2020, the Danish prime minister announced a nationwide lockdown of schools, kindergartens, restaurants, etc. Furthermore, all Danes were encouraged to practice social distancing, and all nonessential employees were encouraged to work from home. Following a marked decrease in infection rates, the lockdown restrictions were gradually lifted after approximately four weeks.

### Participants

We invited 6,000 adult (≥18 years old) patients from the psychiatric services of the Central Denmark Region to participate. Eligible patients were in treatment at the time of the data extraction and had at least one psychiatric contact (inpatient or outpatient) in the three months (91 days) leading up to June 25, 2020. Patients sentenced to treatment (forensic sanction) or diagnosed with an organic mental disorder or mental retardation (International Classification of Diseases [ICD-10] codes F0x.x and F7x.x, respectively) were ineligible. After conducting the survey, we further restricted the population to patients who had been diagnosed with a mental illness (ICD-10 codes: F1x–F9x) prior to the nationwide lockdown on March 11, 2020.

### Survey procedure and questionnaire

The invitation to participate was sent to 6,000 randomly drawn patients via the electronic mailing system used by the Danish authorities (e-Boks).^25^ After describing the aim of the study, the invitation included a personal link to the online survey (an English translation of the questionnaire is available in Supplement 1). The first part of the questionnaire consisted of six questions pertaining to sociodemographics (household size, country of birth, educational level, and occupational status). The second part of the questionnaire was the five-item World Health Organization Well-Being Index (WHO-5), which yields a total score between 0 and 100 where higher scores indicate greater psychological well-being. The third part of the questionnaire was the 18-item Brief Symptom Inventory (BSI-18),^26^ which is a shortened version of the Symptom Checklist-90^27,28^ that contains three six-item subscales: somatization, depression, and anxiety. The total score on the BSI-18 ranges from 0 to 72 with higher scores indicating greater symptom levels. One BSI-18 question “Thoughts of ending your life?” was replaced with another Symptom Checklist-90 question, namely “Thoughts of death or dying?” for ethical reasons (researchers would not be able to act immediately on suicidal intents or plans). The fourth part of the questionnaire consisted of 14 questions exploring changes in mental health and compliance to treatment during the COVID-19 lockdown. This part included a brief summary of the lockdown and its restrictions followed by questions regarding overall mental health; sleep and appetite disturbances; self-harm; substance abuse; medication compliance; and symptoms of anxiety, depression, obsessions and compulsions, somatization, mania, psychosis, eating disorders and negative symptoms. The participants were instructed to rate each of the 14 questions on a five-point Likert-type scale (“Much worse,” “Slightly worse,” “Approximately the same,” “Slightly better,” “Much better”) using the pre-pandemic period as reference. Each question included a brief description of the symptom/construct of interest (e.g., “Depression: Symptoms of depression include feelings of sadness, guilt, decreased pleasure or interest in things and/or people, lack of energy, increased tiredness, reduced self-esteem, self-blame, thoughts of death, trouble concentrating, sleep disturbances, and weight changes”). After answering these 14 questions, respondents who had reported a worsening in their overall mental health during the lockdown (those answering either “Much worse” or “Slightly worse” to the question: “How do you consider your mental health during the lockdown compared to the period before the coronavirus pandemic came to Denmark?” were asked to report perceived reasons for this deterioration from a list of options (available in Supplement 1). The survey was distributed on June 30, 2020, and a reminder was sent to non-respondents on July 7, 2020. The survey closed on July 20, 2020.

### Supplementary data from electronic health records

Sociodemographic (age, sex, civil status, and municipality of residence) and clinical data (most recent main diagnosis [according to the ICD-10], duration of potential admissions, and number of outpatient contacts for the five years prior to June 25, 2020) were extracted from the electronic medical records of all invitees. In defining diagnostic subgroups, the following ICD-10 code hierarchy was employed based on the patients’ most recent main diagnosis: F2x (psychotic disorders) > F3x (mood disorders) > F4x (anxiety- and stress-related disorders) > F5x (eating, sleeping, and other behavioral syndromes associated with physiological disturbances) > F6x (personality disorders) > F8x (developmental disorders including autism) > F9x (child and adolescent mental disorders) > F1x (substance abuse disorders).

### Legal and ethical considerations

This study was approved by the Danish Patient Safety Authority, the Legal Office of the Central Denmark Region, and the medical directors of the hospitals in the Central Denmark Region. All data were processed and stored according to the European Union General Data Protection Regulation. Ethical review board approval is not required for survey-based studies in Denmark. The participants were encouraged to contact the psychiatric services (a hotline number was provided) or their general practitioner if they experienced symptom exacerbation.

### Statistical analysis

Demographic and clinical variables are presented as numbers and percentages for categorical variables and means and standard variations (SD) for continuous variables (medians and interquartile ranges [IQR] for non-normally distributed data). Fisher’s exact test and Student’s simple t-test were applied to compare categorical and continuous estimates between respondents and non-respondents. To minimize the effect of selection bias, we weighted estimates of the outcomes (the WHO-5 total score, the BSI-18 total and subscale scores, and the proportions of respondents in the response categories for the 14 questions regarding changes in mental health during the lockdown) using the inverse propensity weighted (IPW) estimation method.^29^ The IPW method weighted these estimates according to the demographic and clinical variables on which the respondents and non-respondents differed with statistical significance, namely sex, civil status, municipality, diagnosis, and number of pre-pandemic outpatient visits. To determine potential risk factors for mental health deterioration, we performed a logistic regression with the weighted response to the question regarding mental health deterioration during lockdown as the outcome (respondents answering either “Much worse” or “Slightly worse” to the question: “How do you consider your mental health during the lockdown compared to the period before the coronavirus pandemic came to Denmark?”) and demographic and clinical characteristics as explanatory variables. All analyses were performed using the R statistical software version 4.0.1. The significance level was set at .05.

## RESULTS

### Sample characteristics

Figure 1 illustrates the sampling and recruitment procedure. A total of 992 individuals with mental illness diagnosed prior to the Spring 2020 lockdown responded to the questionnaire (response rate of 21.6%).

**Figure 1:**
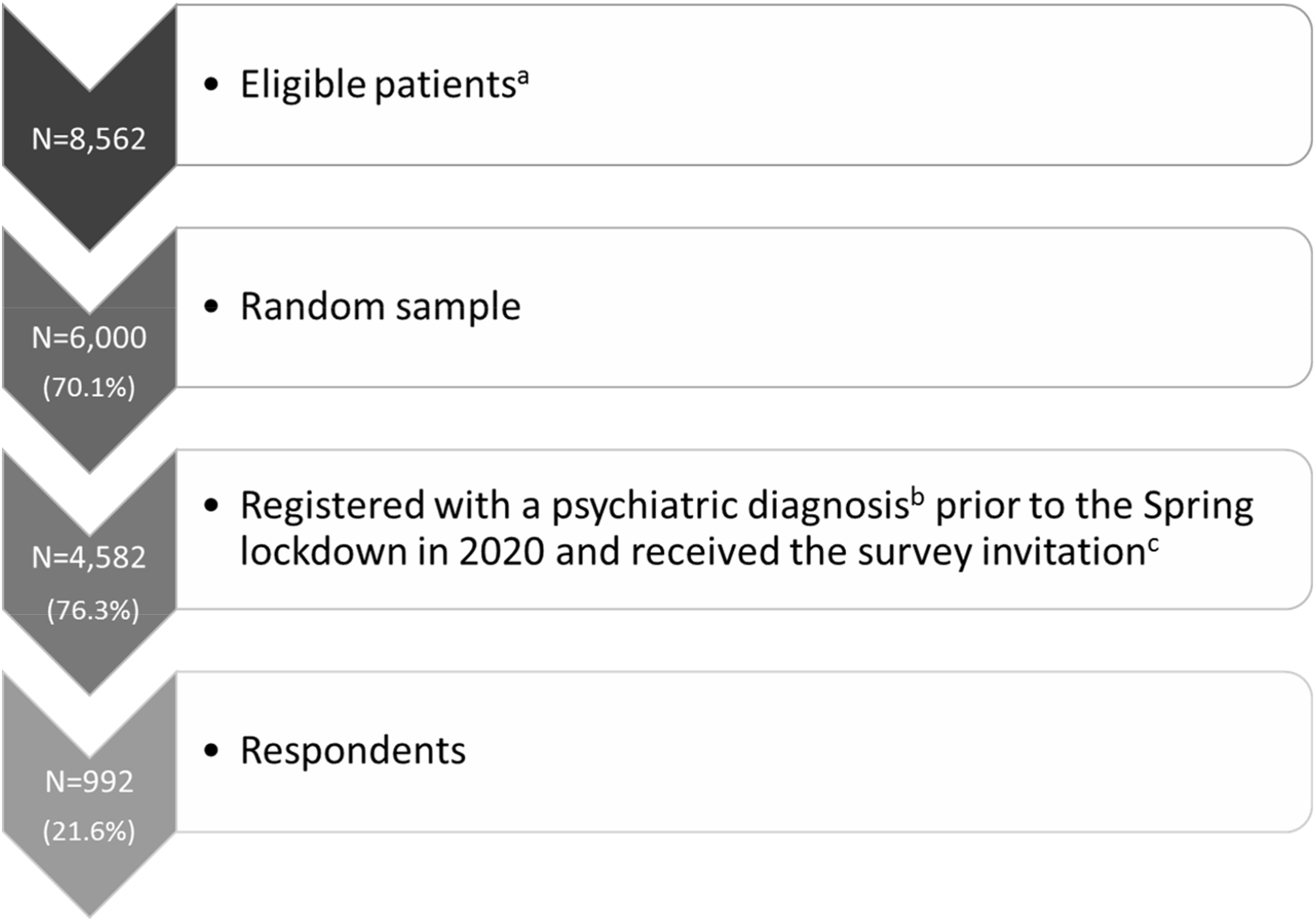
Flowchart of the study population including response rate for the questionnaire. ^**a**^ Eligibility criteria: patients from Psychiatric Services of Central Denmark Region, ≥18 years old, having at least one psychiatric contact in the three months (91) days before June 25, 2020. Patients with forensic sanction or diagnosed with an organic mental disorder or mental retardation were not eligible. ^**b**^ A clinical diagnosis of F2x (psychotic disorders), F3x (mood disorders), F4x (anxiety- and stress-related disorders), F5x (eating-, sleeping- and other behavioral syndromes associated with physiological disturbances), F6x (personality disorders), F8x (developmental disorders incl. autism), F90-F98 (child- and adolescent mental disorders, or F1x (substance abuse disorders) prior to the nationwide lockdown (March 11, 2020). ^**c**^ Invitation were sent via the electronic mailing system (e-Boks) used by the Danish authorities.

Table 1 lists the sociodemographic and clinical characteristics of respondents and non-respondents. Compared to non-respondents, respondents were more likely to be female and married, to live in a municipality with >150.000 inhabitants, have a diagnosis of depression, and have attended more outpatient visits both before and during the pandemic. The majority of respondents were born in Denmark (91.6%) and lived with at least one other adult (60.6%). A relatively large proportion of the respondents were absent owing to illness (35.8%).

**Table 1:**
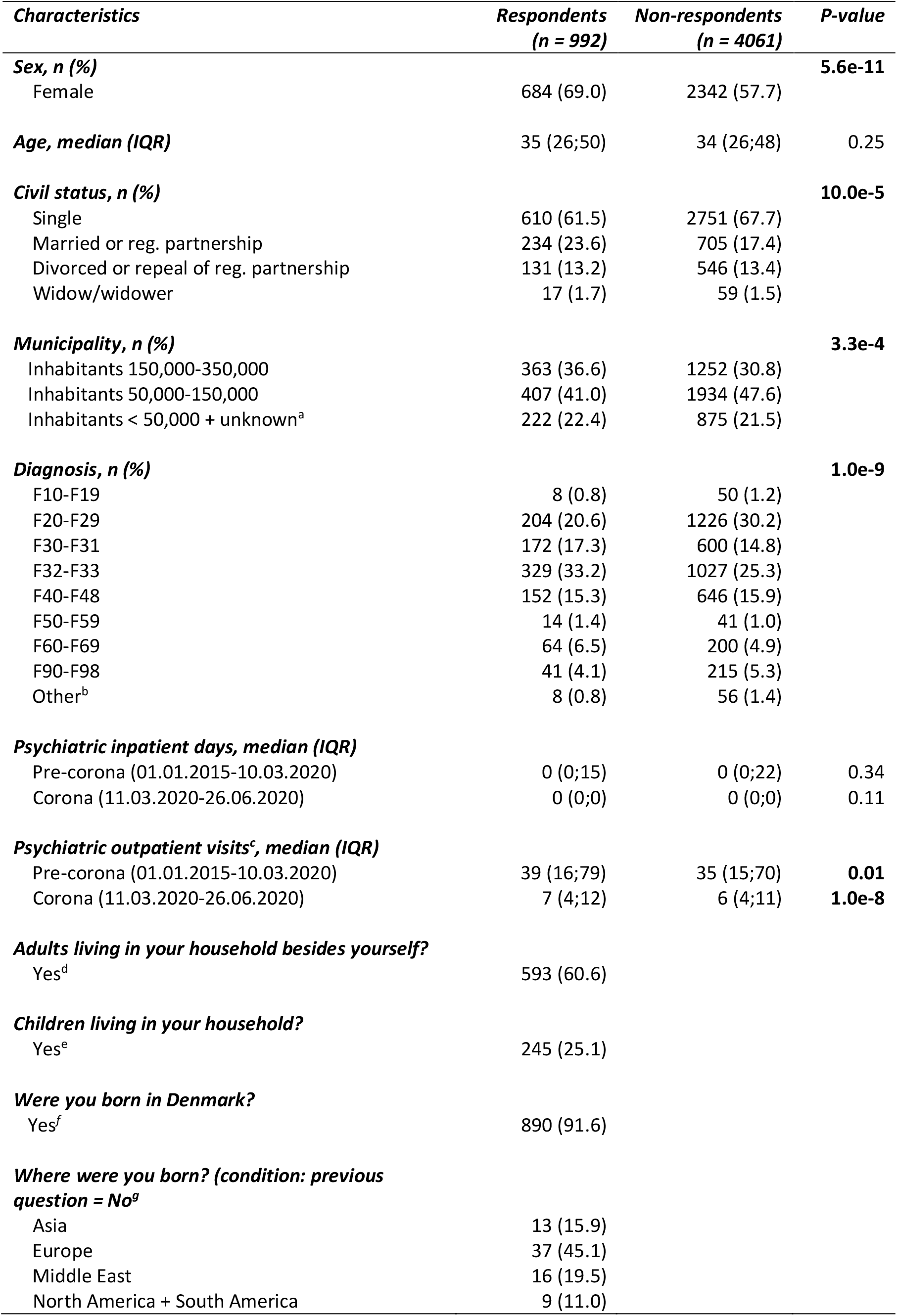
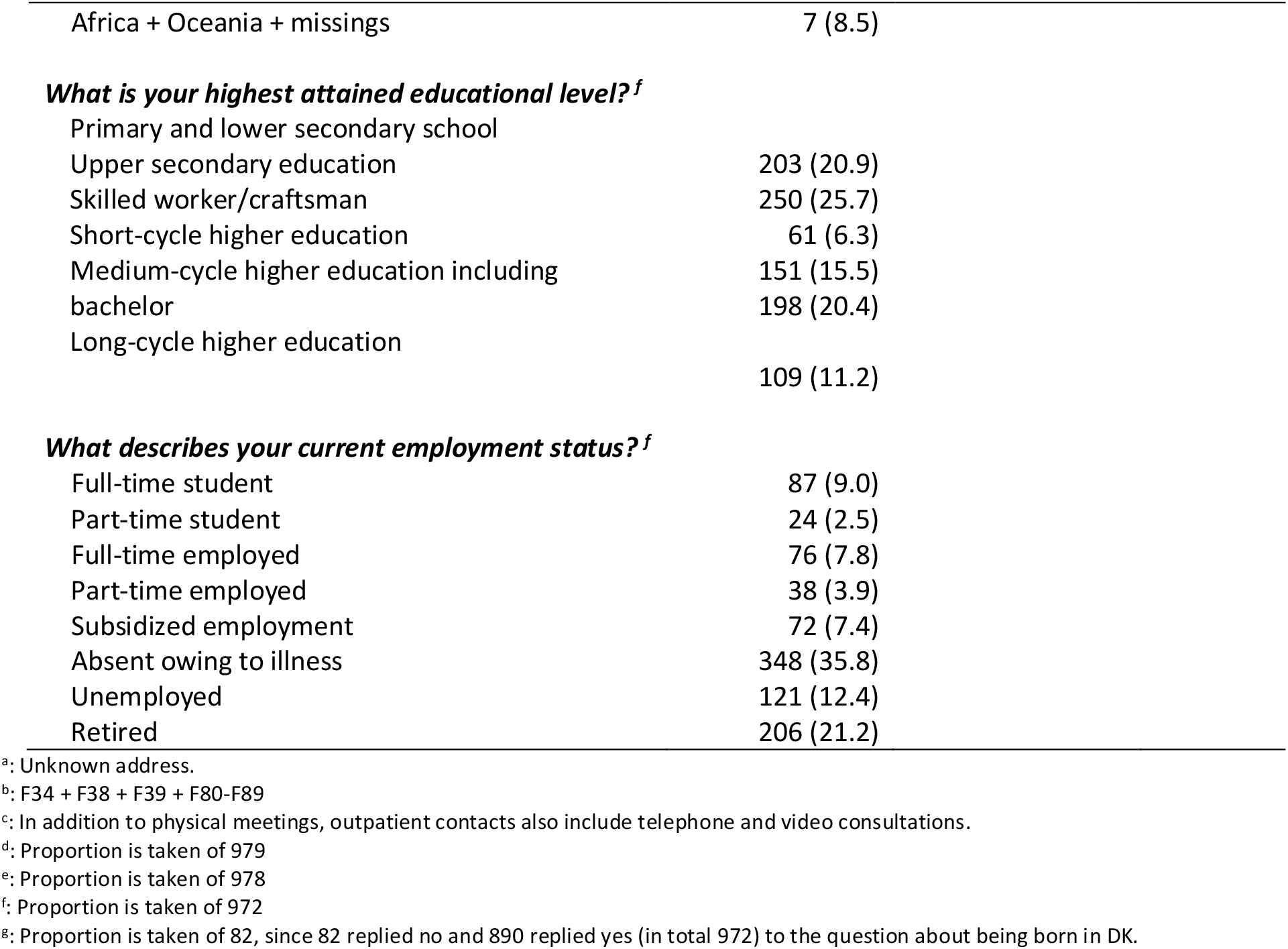
Sociodemographic and clinical characteristics of respondents versus non-respondents. The p-values are based upon t-test for the continuous values and Fisher’s exact test or chi-square test for the categorical variables. Significant p-values are marked in **bold**.

### Weighting of results based on attrition

The differences between the weighted and non-weighted means of the WHO-5 total score, the BSI-18 total and subscale scores, and responses to the 14 mental health questions were very minor, and none were statistically significant (see Supplement 2).

### Self-reported mental health

The mean weighted WHO-5 total score was 38.1 (SD=22.7), while the mean BSI-18 total score was 28.4 (SD=15.2) with mean scores of 6.4 (5.0), 11.9 (SD=6.3), and 10.1 (SD=5.8) on the somatization, depression and anxiety subscales, respectively. The WHO-5 and BSI-18 scores for the following diagnostic groups— psychotic disorder (ICD-10: F2x), bipolar disorder (ICD-10: F30–31), unipolar depression (ICD-10: F32–33), anxiety disorder (ICD-10: F4x), and personality disorder (ICD-10: F6x)—are available in Supplement 2. WHO-5 total scores ranged from 34.1 (SD=20.9) for respondents with unipolar depression to 42.9 (23.1) for those with psychotic disorder, and BSI-18 total scores ranged from 25.6 (SD=14.8) for those with psychotic disorder to 33.1 (SD=16.7) for those with anxiety disorder.

### Changes in mental health during lockdown

Figure 2 shows the response to the questions assessing changes in mental health during the Spring lockdown. A total of 52% of respondents reported their overall mental health had deteriorated during the lockdown, while 33% reported no change and 16% reported improvement. The pattern was equivalent for specific symptom domains (predominantly worsening). Across the diagnostic groups, the proportion of participants reporting deterioration of mental health ranged from 44% among those with psychotic disorder to 61% among those with personality disorder.

**Figure 2:**
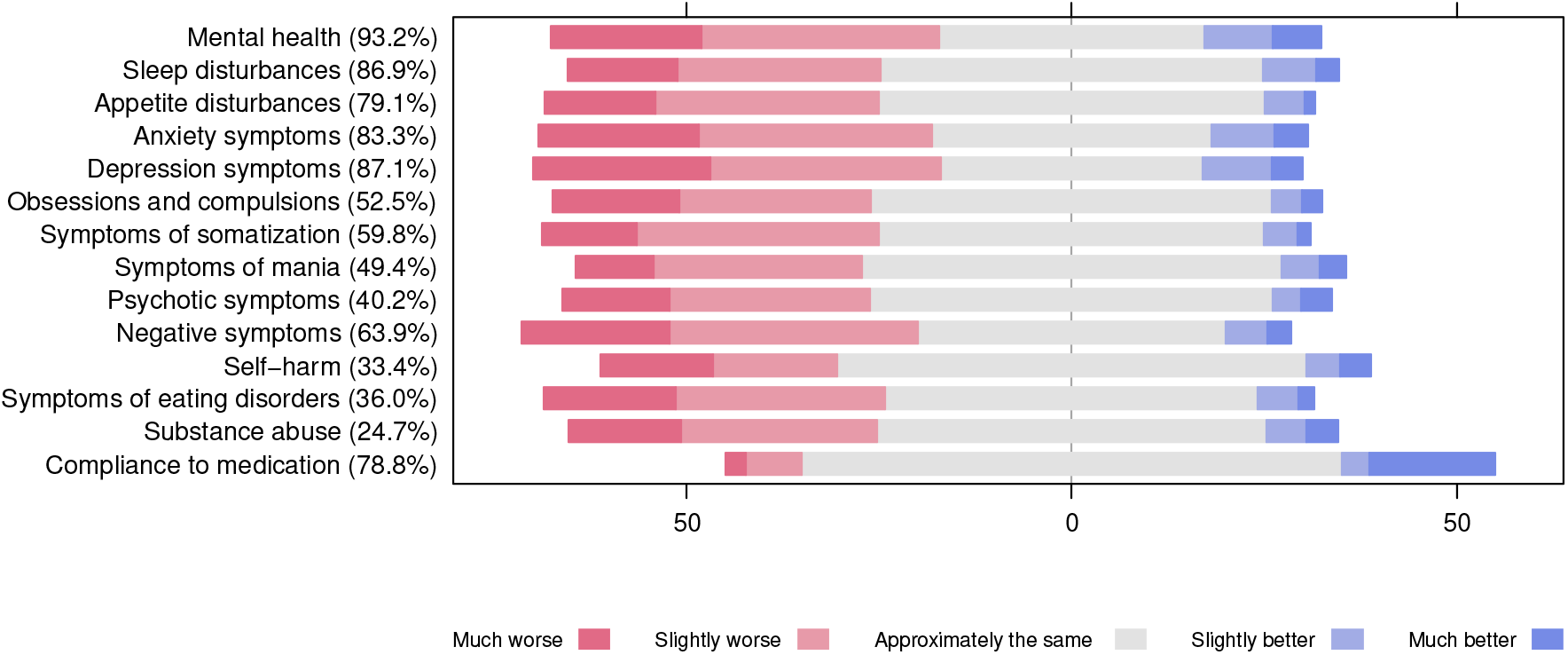
Weighted response to the 14 questions focusing on changes in mental health and compliance to treatment during the COVID-19 lockdown of Denmark in the spring of 2020. The numbers in the parentheses represent the proportion of respondents endorsing one of the five likert-scale responses (they could also choose: “The symptom was not present (either before or during the lockdown)” or choose not to respond). Equivalent results stratified on the following diagnostic groups: psychotic disorder (ICD-10: F2x), bipolar disorder (ICD-10: F30-31), unipolar depression (ICD-10: F32-33), anxiety disorder (ICD-10: F4x), and personality disorder (ICD-10: F6x) are available in Supplementary Figures 3-7 and Supplementary Table 3.

### Characteristics associated with worsening of mental health during lockdown

Table 2 presents the results of the logistic regression analysis of the association between patient characteristics and reported worsening of overall mental health during lockdown. The only characteristic positively associated with mental health deterioration during lockdown was living with no other adults in the household (odds ratio for no other adult versus at least one other adult in the household: 1.41 [95%CI: 1.02;1.94], p = 0.038). A main diagnosis of psychotic disorder (ICD-10: F2x) was negatively associated with mental health deterioration during lockdown (odds ratio for psychotic disorder versus unipolar depression: 0.56 [95%CI: 0.36;0.85], p = 0.007).

**Table 2:**
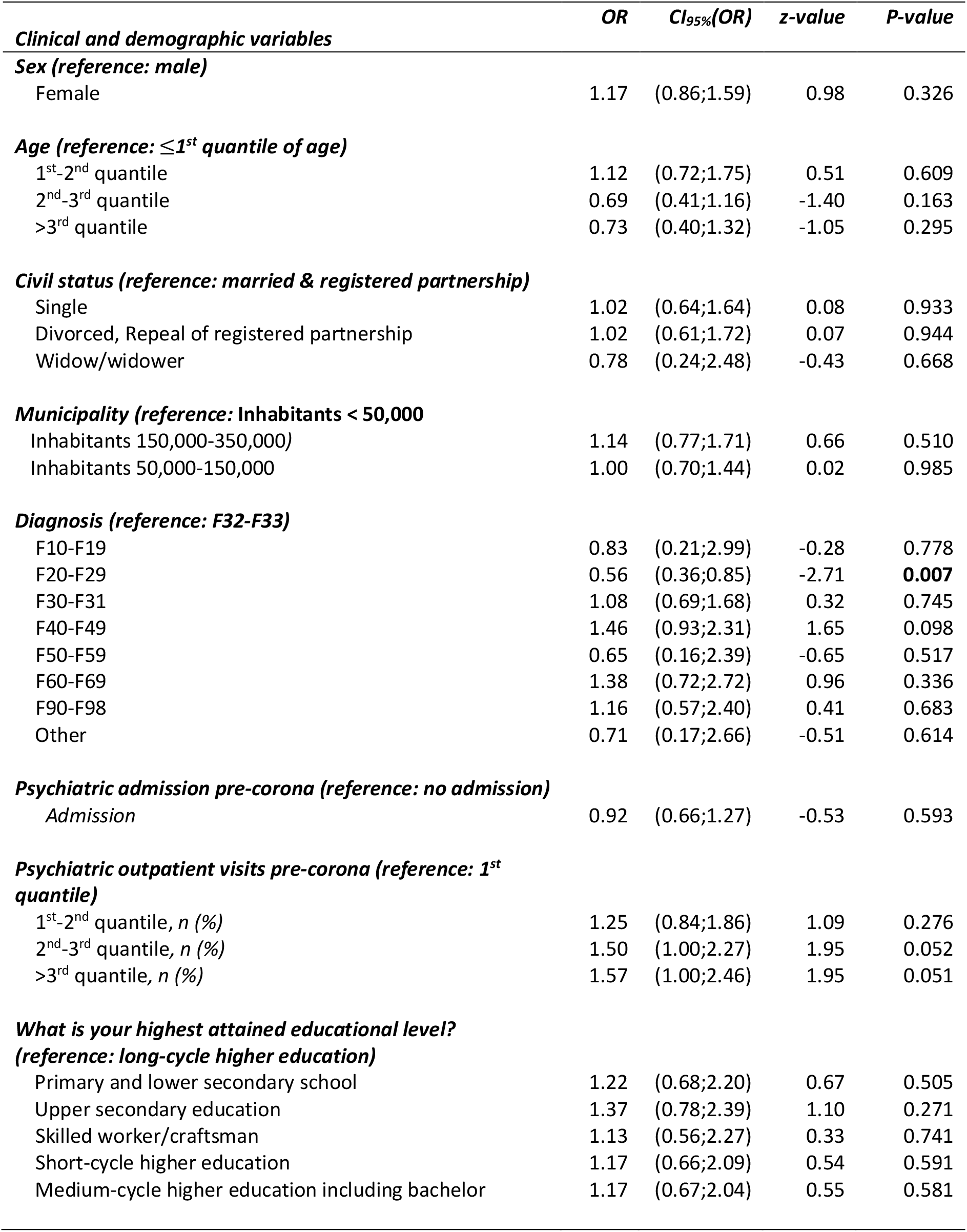
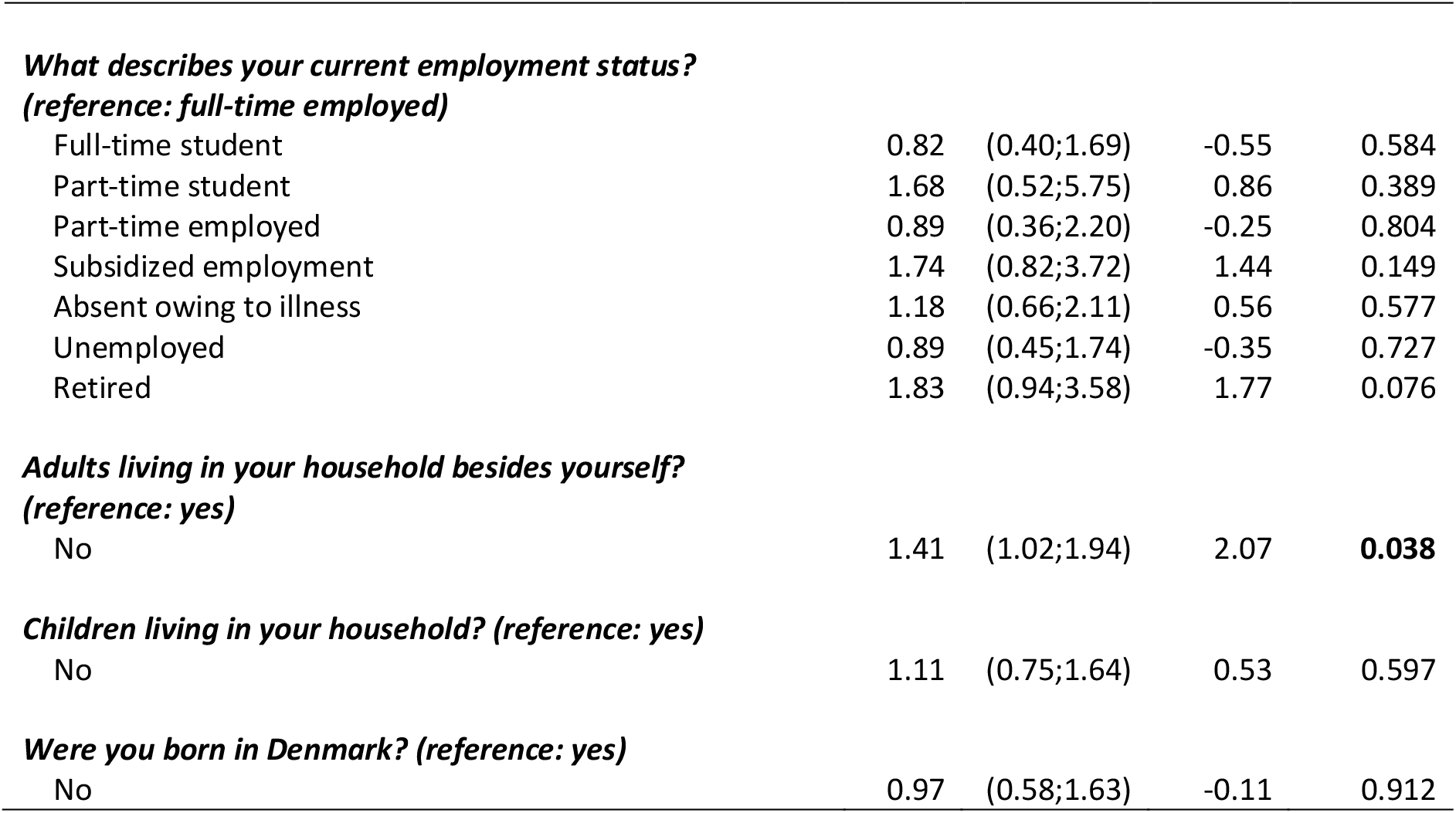
Logistic regression with weighted deterioration in mental health as outcome (participants responding either ‘Much worse’ or ‘Slightly worse’ to the question: “How do you consider your mental health during the lockdown compared to the period before the corona pandemic came to Denmark?”).

|  | *OR* | *CI<sub>95%</sub>(OR)* | *z-value* | *P-value* |
| --- | --- | --- | --- | --- |
| ***Clinical and demographic variables*** |  |  |  |  |
| ***Sex (reference: male)*** |  |  |  |  |
| Female | 1.17 | (0.86;1.59) | 0.98 | 0.326 |
| ***Age (reference: ≤1<sup>st</sup> quantile of age)*** |  |  |  |  |
| 1<sup>st</sup>-2<sup>nd</sup> quantile | 1.12 | (0.72;1.75) | 0.51 | 0.609 |
| 2<sup>nd</sup>-3<sup>rd</sup> quantile | 0.69 | (0.41;1.16) | -1.40 | 0.163 |
| >3<sup>rd</sup> quantile | 0.73 | (0.40;1.32) | -1.05 | 0.295 |
| ***Civil status (reference: married & registered partnership)*** |  |  |  |  |
| Single | 1.02 | (0.64;1.64) | 0.08 | 0.933 |
| Divorced, Repeal of registered partnership | 1.02 | (0.61;1.72) | 0.07 | 0.944 |
| Widow/widower | 0.78 | (0.24;2.48) | -0.43 | 0.668 |
| ***Municipality (reference: Inhabitants < 50,000)*** |  |  |  |  |
| Inhabitants 150,000-350,000) | 1.14 | (0.77;1.71) | 0.66 | 0.510 |
| Inhabitants 50,000-150,000 | 1.00 | (0.70;1.44) | 0.02 | 0.985 |
| ***Diagnosis (reference: F32-F33)*** |  |  |  |  |
| F10-F19 | 0.83 | (0.21;2.99) | -0.28 | 0.778 |
| F20-F29 | 0.56 | (0.36;0.85) | -2.71 | **0.007** |
| F30-F31 | 1.08 | (0.69;1.68) | 0.32 | 0.745 |
| F40-F49 | 1.46 | (0.93;2.31) | 1.65 | 0.098 |
| F50-F59 | 0.65 | (0.16;2.39) | -0.65 | 0.517 |
| F60-F69 | 1.38 | (0.72;2.72) | 0.96 | 0.336 |
| F90-F98 | 1.16 | (0.57;2.40) | 0.41 | 0.683 |
| Other | 0.71 | (0.17;2.66) | -0.51 | 0.614 |
| ***Psychiatric admission pre-corona (reference: no admission)*** |  |  |  |  |
| Admission | 0.92 | (0.66;1.27) | -0.53 | 0.593 |
| ***Psychiatric outpatient visits pre-corona (reference: 1<sup>st</sup> quantile)*** |  |  |  |  |
| 1<sup>st</sup>-2<sup>nd</sup> quantile, *n* (%) | 1.25 | (0.84;1.86) | 1.09 | 0.276 |
| 2<sup>nd</sup>-3<sup>rd</sup> quantile, *n* (%) | 1.50 | (1.00;2.27) | 1.95 | 0.052 |
| >3<sup>rd</sup> quantile, *n* (%) | 1.57 | (1.00;2.46) | 1.95 | 0.051 |
| ***What is your highest attained educational level? (reference: long-cycle higher education)*** |  |  |  |  |
| Primary and lower secondary school | 1.22 | (0.68;2.20) | 0.67 | 0.505 |
| Upper secondary education | 1.37 | (0.78;2.39) | 1.10 | 0.271 |
| Skilled worker/craftsman | 1.13 | (0.56;2.27) | 0.33 | 0.741 |
| Short-cycle higher education | 1.17 | (0.66;2.09) | 0.54 | 0.591 |
| Medium-cycle higher education including bachelor | 1.17 | (0.67;2.04) | 0.55 | 0.581 |
| ***What describes your current employment status?*** |  |  |  |  |
| ***(reference: full-time employed)*** |  |  |  |  |
| Full-time student | 0.82 | (0.40;1.69) | -0.55 | 0.584 |
| Part-time student | 1.68 | (0.52;5.75) | 0.86 | 0.389 |
| Part-time employed | 0.89 | (0.36;2.20) | -0.25 | 0.804 |
| Subsidized employment | 1.74 | (0.82;3.72) | 1.44 | 0.149 |
| Absent owing to illness | 1.18 | (0.66;2.11) | 0.56 | 0.577 |
| Unemployed | 0.89 | (0.45;1.74) | -0.35 | 0.727 |
| Retired | 1.83 | (0.94;3.58) | 1.77 | 0.076 |
| ***Adults living in your household besides yourself?*** |  |  |  |  |
| ***(reference: yes)*** |  |  |  |  |
| No | 1.41 | (1.02;1.94) | 2.07 | **0.038** |
| ***Children living in your household? (reference: yes)*** |  |  |  |  |
| No | 1.11 | (0.75;1.64) | 0.53 | 0.597 |
| ***Were you born in Denmark? (reference: yes)*** |  |  |  |  |
| No | 0.97 | (0.58;1.63) | -0.11 | 0.912 |

### Reasons for worsening of mental health

Figure 3 shows the perceived reasons for worsening among the 52% of respondents who reported that their overall mental health had deteriorated during lockdown. The most commonly reported reasons for deterioration were “loneliness” (67%), “disruption of routines” (66%), “concerns about family/friends getting infected with coronavirus” (59%), “less contact with friends” (59%), “less contact with family” (51%), “concerns about getting infected with coronavirus” (46%), “boredom” (43%), and “reduced access to psychiatric treatment” (43%).

**Figure 3:**
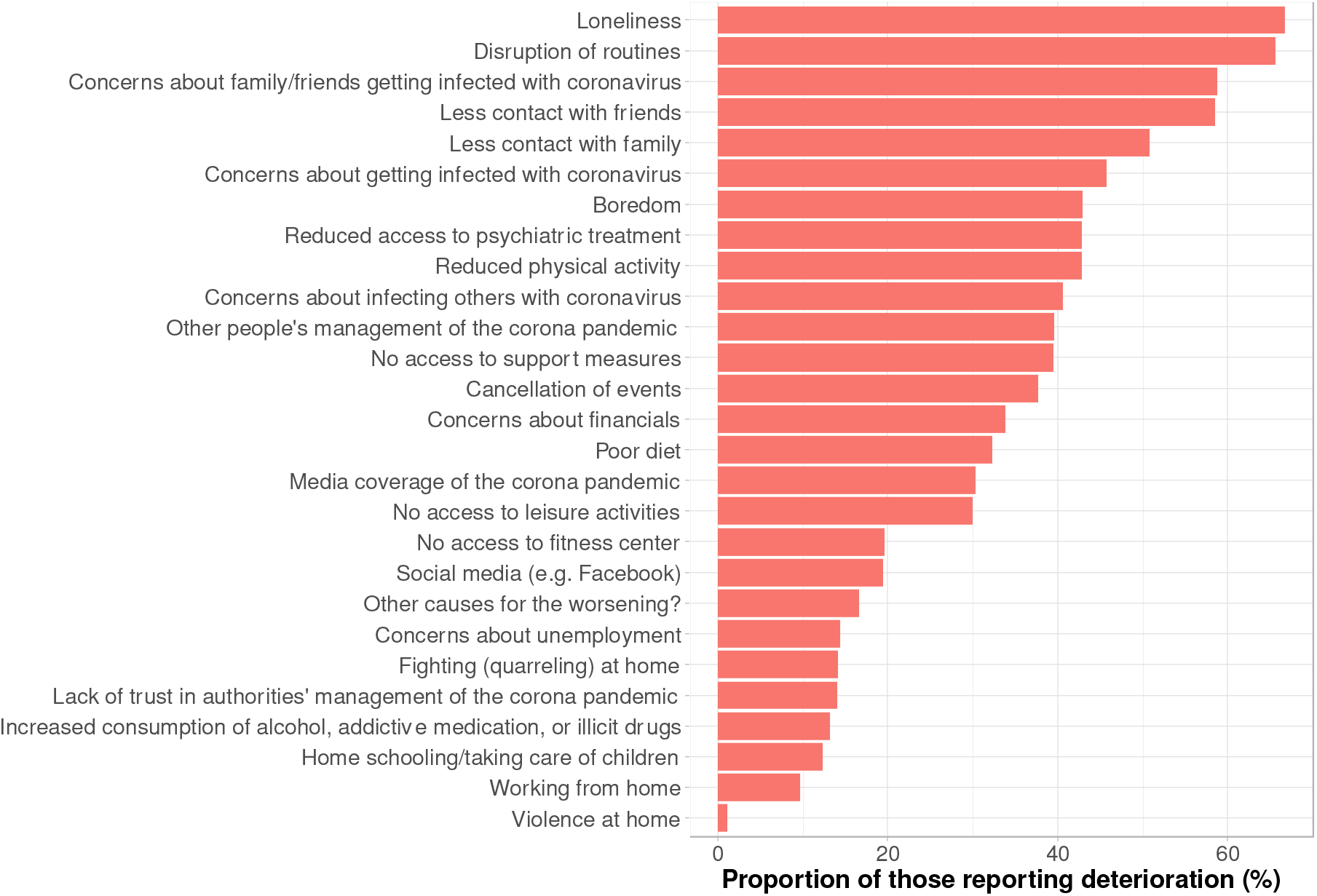
Perceived reasons for deterioration during the pandemic lockdown (weighted). Perceived reasons for deterioration among the 479/925 (52%) of the respondents who reported that their overall mental health had worsened during the nationwide pandemic lockdown stratified on the following diagnostic groups: psychotic disorder (ICD-10: F2x), bipolar disorder (ICD-10: F30-31), unipolar depression (ICD-10: F32-33), anxiety disorder (ICD-10: F4x), and personality disorder (ICD-10: F6x) are available in Supplementary Figure 2 and Supplementary Table 2.

The above results are based on the population restricted to patients who had been diagnosed with a mental illness (ICD-10 codes: F1x–F9x) prior to the nationwide lockdown on March 11, 2020. Results on the full population i.e. including patients with a Zx code (contact with psychiatric health services incl. examination) prior to the nationwide lockdown are available in Supplement 3.

## DISCUSSION

In this survey of patients with mental illness, more than half of respondents reported deteriorating mental health during the nationwide pandemic lockdown of Denmark in the Spring of 2020, while only 16% reported improvement. Living with no other adults in the household was the only statistically significant risk factor for reported worsening in mental health during the lockdown. The respondents reported loneliness, disruption of routines, concerns about family/friends/self becoming infected with coronavirus, less frequent contact with family/friends, boredom, and reduced access to psychiatric treatment as the most common reasons for the deterioration in their mental health.

As expected for a sample of patients with mental illness under psychiatric treatment, the level of self-reported symptoms was high, while the level of psychological well-being was low. Specifically, respondents reported symptom severity (somatization, depression, and anxiety) on the BSI-18 similar to^30^ or slightly higher than^31,32^ those previously reported for psychiatric outpatients. Accordingly, the level of psychological well-being measured by the WHO-5 (mean total score: 38) aligned with those found in studies of psychiatric outpatients,^33^ and was considerably lower than that of the general Danish population during the pandemic (mean WHO-5 total scores: 62–65).^3,5^

As in other cross-sectional studies, a large proportion of the respondents with mental illness (52%) reported worsening in their overall mental health during the pandemic lockdown.^8-16^ Conversely, several longitudinal studies have reported either no change in psychiatric symptoms^6,17-19^ or even an improvement ^14,20,21^ among psychiatric patients during the pandemic. However, as the authors of one of the studies reporting improvement explained,^14^ this finding might be attributable to selection bias as non-respondents were more likely than respondents to have pre-existing mental illness and low educational attainment. We aimed at minimizing this potential bias via random sampling and weighting of response based on attrition. Notably, we found no differences between weighted and non-weighted results, which corroborates the validity of results from other studies that employed random sampling in clinical populations, but included no or limited data on non-respondents.^6,21^

Understanding the mechanisms underlying the negative impact of the COVID-19 pandemic on mental health is important to inform measures to alleviate it. In the present study, living with no other adults in the household was the characteristic that was positively associated (with statistical significance) to deterioration of mental health during the pandemic lockdown. This finding is consistent with the most common reasons respondents offered for their deteriorating mental health, namely loneliness and less frequent contact with friends and family. Disruption of routines and reduced access to psychiatric care were other very common self-reported reasons for deterioration in mental health, both of which seem intuitively meaningful and are consistent with findings from other studies.^9,34^ While these findings should ideally be replicated in other settings, they offer a cautious recommendation for an increased emphasis on ensuring social and clinical support for individuals with mental illness during the ongoing and potential future pandemics.^1,35,36^

### Limitations

There are limitations to this study, which should be taken into account by the reader. First, the response rate was rather low although similar to that of other surveys among individuals with mental disorders.^6,8^ However, a main strength of the study was the possibility to weigh the results based on attrition, which will have reduced the impact of potential selection bias. To further reduce selection bias, invitation to the survey was based on random sampling. Second, the validity of self-reported symptoms and psychological well-being may be questionable if respondents have poor insight or reduced cognitive function. However, self-reported and clinician-rated psychiatric symptoms generally exhibit high concordance,^37,38^ which supports the validity of the findings of this study. Third, invitations to participate were distributed 11 weeks after the lockdown was lifted, which could have led to recall bias. To reduce the impact of this potential bias, we included a brief summary of the lockdown’s events and restrictions in the questionnaire. Relatedly, patients’ general attitudes could influence their perceptions of the reasons for the deterioration of their mental health and the reported reasons may therefore not necessarily reflect the impact of the lockdown per se. For instance, patients who were generally unsatisfied with their psychiatric treatment may have consciously or subconsciously used the survey as an opportunity to voice this opinion and thus have attributed their perceived deterioration to “poorer access to psychiatric treatment.” However, psychiatric care has objectively changed during the pandemic,^34,39-41^ and it is concerning that nearly 43% of the respondents who reported an overall worsening of their mental health during lockdown attributed it, at least in part, to reduced access to treatment. Fifth, the findings of the present study may not generalize to other countries and health care settings. Compared to many other countries, the first wave of the pandemic took a relatively benign course in Denmark.^42^ Hence, the lockdown’s apparent impact on Danish patients with mental illness may have been equally benign compared to that experienced in countries where the pandemic was more severe. Sixth, this study was cross-sectional with retrospective assessment of the lockdown. Ideally, future studies with random sampling and weighting for attrition should evaluate the long-term effects of the COVID-19 pandemic upon individuals with mental illness.

In this survey of patients with mental disorders, more than half of respondents reported a worsening of their mental health during the pandemic lockdown in the Spring of 2020. The most commonly reported reasons for deterioration in mental health were loneliness, disruption of routines, concerns about and less frequent contact with family/friends, boredom, and reduced access to psychiatric care. In addition, living with no other adults in the household was associated with worsening in mental health during the lockdown. These findings suggest that there should be increased emphasis on both social and clinical support measures for individuals with mental illness during the ongoing and potential future pandemics.

## Data Availability

The participants of this study did not agree to their data being shared publicly.

## Acknowledgements

The authors thank Mia Buhl Povlsen and Gitte Kobberøe for practical assistance with the distribution of the questionnaires. Bettina Nørremark is thanked for extraction of data from the medical records. Anders Helles Carlsen is thanked for review of the statistical code.

