## Supplement 3 for "Mental health of patients with mental illness during the COVID-19 pandemic lockdown: A questionnaire-based survey weighted for attrition"

**Figure 1:** Flowchart of the study population including response rate for the questionnaire.

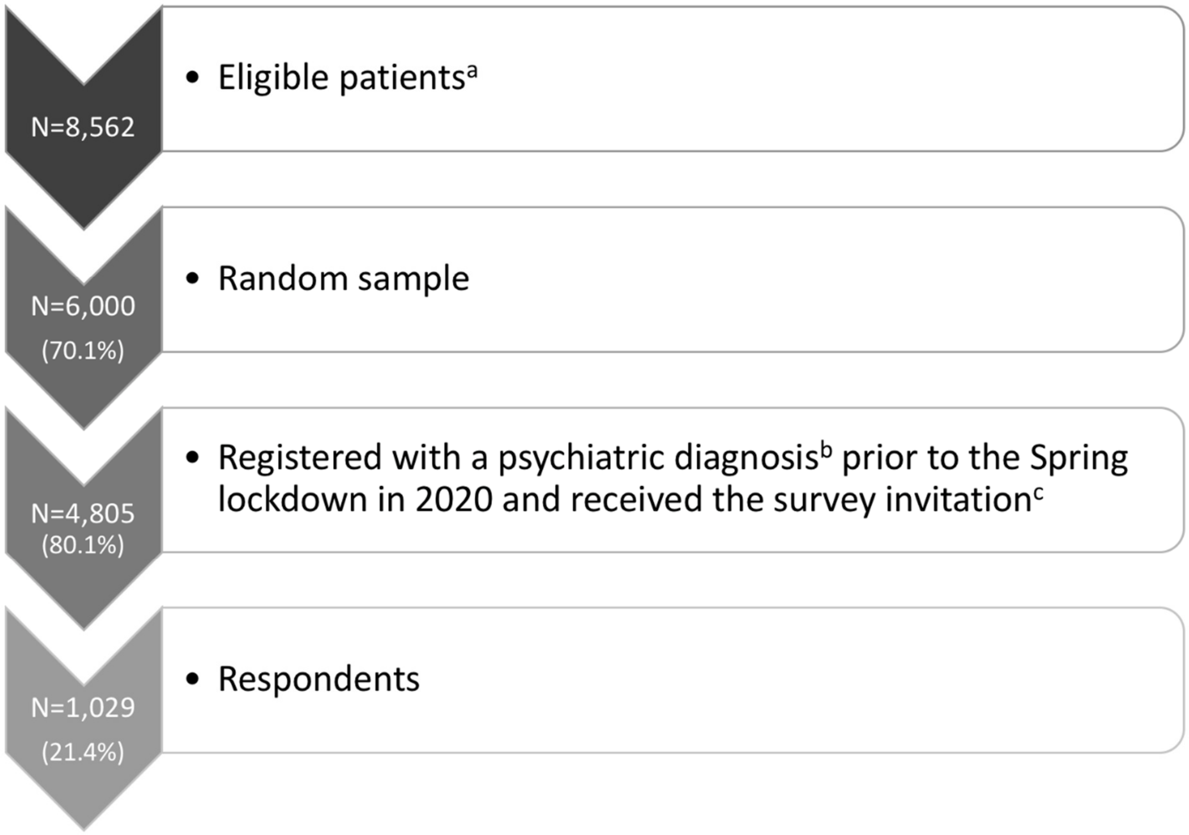

[a] Eligibility criteria: patients from Psychiatric Services of Central Denmark Region, ≥18 years old, having at least one psychiatric contact in the three months (91) days before June 25, 2020. Patients with forensic sanction or diagnosed with an organic mental disorder or mental retardation were not eligible.

[b] A clinical diagnosis of F2x (psychotic disorders), F3x (mood disorders), F4x (anxiety- and stress-related disorders), F5x (eating-, sleeping- and other behavioral syndromes associated with physiological disturbances), F6x (personality disorders), F8x (developmental disorders incl. autism), F90-F98 (child- and adolescent mental disorders, F1x (substance abuse disorders) or Zx (contact with psychiatric health services incl. examination) prior to the nationwide lockdown (March 11, 2020).

[c] Invitation were sent via the electronic mailing system (e-Boks) used by the Danish authorities.

**Table 1:** Sociodemographic and clinical characteristics of respondents versus non-respondents. The p-values are based upon t-test for the continuous values and Fisher's exact test or chi-square test for the categorical variables. Significant p-values are marked in **bold.**

| *Characteristics* | *Respondents (n = 1029)* | *Non-respondents (n = 4258)* | *P-value* |
|---|---|---|---|
| ***Sex, n (%)*** | | | **9.7e-12** |
| Female | 708 (68.8) | 2439 (57.3) | |
| ***Age, median (IQR)*** | 35 (26;50) | 34 (26;48) | 0.25 |
| ***Civil status, n (%)*** | | | **9.5e-5** |
| Single | 638 (62.0) | 2890 (67.9) | |
| Married or reg. partnership | 241 (23.4) | 733 (17.2) | |
| Divorced or repeal of reg. partnership | 133 (12.9) | 572 (13.4) | |
| Widow/widower | 17 (1.7) | 63 (1.5) | |
| ***Municipality, n (%)*** | | | **6.8e-4** |
| Inhabitants 150,000-350,000 | 376 (36.5) | 1322 (31.0) | |
| Inhabitants 50,000-150,000 | 425 (41.3) | 2018 (47.4) | |
| Inhabitants < 50,000 + unknown[a] | 228 (22.2) | 918 (21.6) | |
| ***Diagnosis, n (%)*** | | | **9.5e-10** |
| F10-F19 | 8 (0.8) | 50 (1.2) | |
| F20-F29 | 204 (19.8) | 1226 (28.8) | |
| F30-F31 | 172 (16.7) | 600 (14.1) | |
| F32-F33 | 329 (32.0) | 1027 (24.1) | |
| F40-F48 | 152 (14.8) | 646 (15.2) | |
| F50-F59 | 14 (1.4) | 41 (1.0) | |
| F60-F69 | 64 (6.2) | 200 (4.7) | |
| F90-F98 | 41 (4.0) | 215 (5.0) | |
| Other F-diagnoses[b] | 8 (0.8) | 56 (1.3) | |
| Z-diagnoses | 37(3.6) | 197 (4.6) | |
| ***Psychiatric inpatient days, median (IQR)*** | | | |
| Pre-corona (01.01.2015-10.03.2020) | 0 (0;13) | 0 (0;19) | 0.39 |
| Corona (11.03.2020-26.06.2020) | 0 (0;0) | 0 (0;0) | 0.12 |
| ***Psychiatric outpatient visits[c], median (IQR)*** | | | |
| Pre-corona (01.01.2015-10.03.2020) | 37 (14;77) | 33 (13;67) | **6.2e-3** |
| Corona (11.03.2020-26.06.2020) | 7 (4;12) | 6 (4;10) | **2.0e-9** |
| ***Adults living in your household besides yourself?*** | | | |
| Yes[d] | 617 (60.8) | | |
| ***Children living in your household?*** | | | |
| Yes[e] | 250 (24.7) | | |
| ***Were you born in Denmark?*** | | | |
| Yes[f] | 919 (91.2) | | |
| ***Where were you born? (condition: previous question = No[g])*** | | | |
| Asia | 14 (15.7) | | |
| Europe | 42 (47.2) | | |
| Middle East | 16 (18.0) | | |

| | |
|---|---|
| North America + South America | 10 (11.2) |
| Africa + Oceania + unknown | 7 (7.9) |
| ***What is your highest attained educational level?*** [f] | |
| Primary and lower secondary school | 214 (21.2) |
| Upper secondary education | 258 (25.6) |
| Skilled worker/craftsman | 64 (6.3) |
| Short-cycle higher education | 157 (15.6) |
| Medium-cycle higher education including bachelor | 206 (20.4) |
| Long-cycle higher education | 109 (10.8) |
| ***What describes your current employment status?*** [f] | |
| Full-time student | 92 (9.1) |
| Part-time student | 27 (2.7) |
| Full-time employed | 83 (8.2) |
| Part-time employed | 39 (3.9) |
| Subsidized employment | 73 (7.2) |
| Absent owing to illness | 359 (35.6) |
| Unemployed | 125 (12.4) |
| Retired | 210 (20.8) |

[a]: Unknown address.
[b]: F34 + F38 + F39 + F80-F89
[c]: In addition to physical meetings, outpatient contacts also include telephone and video consultations.
[d]: Proportion is taken of 1015
[e]: Proportion is taken of 1012
[f]: Proportion is taken of 1008
[g]: Proportion is taken of 89, since 89 replied no and 919 replied yes (in total 1008) to the question about being born in DK.

**Figure 2:** Visualization of the non-weighted and weighted mean and 95%-confidence interval of the WHO-5 total score, the BSI-18 total score, the three subscales of BSI-18.

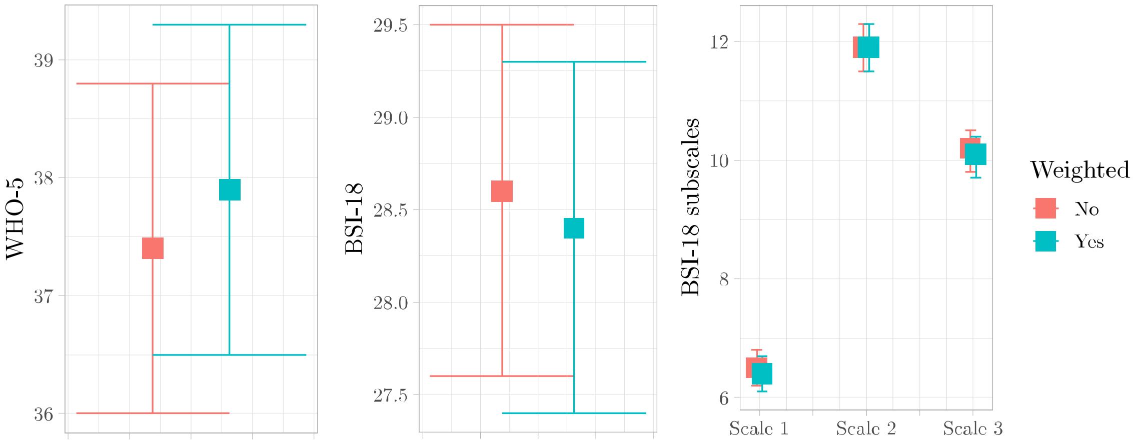

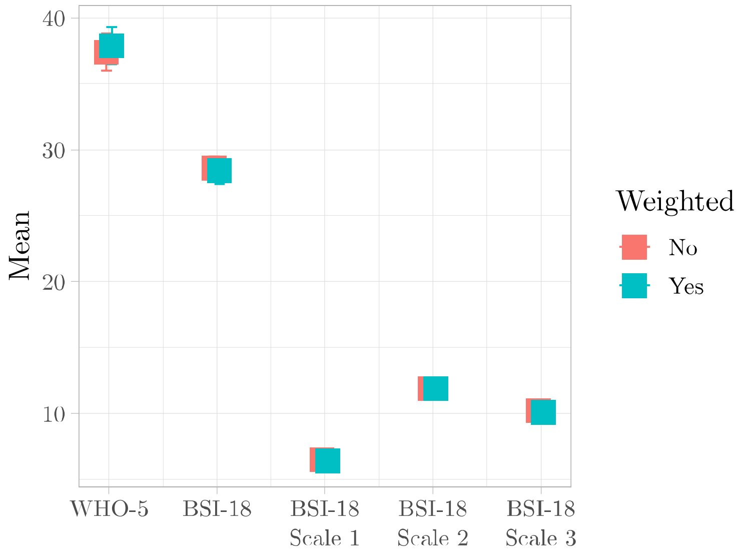

**Table 2:** Table displaying the non-weighted (red) and weighted (blue) mean (SD) of the WHO-5 total score, the BSI-18 total score, the three subscales of BSI-18 for all respondents and stratified on the ICD-10 diagnosis: F2, F30-F31, F32-F33, F4 and F6.

| ***Questionnaires*** *Mean (SD)* | ***Respondents*** *(n = 992)* | ***F2*** *(n = 204)* | ***F30-F31*** *(n = 172)* | ***F32-F33*** *(n = 329)* | ***F4*** *(n = 152)* | ***F6*** *(n = 64)* |
|---|---|---|---|---|---|---|
| ***WHO-5*** | | | | | | |
| Total score* | 37.4 (22.4) | 42.9 (23.3) | 38.6 (23.5) | 34.1 (20.8) | 35.8 (22.1) | 34.9 (19.8) |
| | 37.9 (22.6) | 42.9 (23.1) | 38.5 (23.6) | 34.2 (20.9) | 35.2 (22.1) | 35.0 (20.0) |
| ***BSI-18*** | | | | | | |
| Total score | 28.6 (14.9) | 25.9 (14.9) | 25.5 (15.5) | 29.5 (13.4) | 32.7 (16.9) | 31.0 (13.6) |
| | 28.4 (15.1) | 25.6 (14.8) | 25.9 (15.9) | 29.4 (13.5) | 33.0 (16.7) | 31.0 (13.4) |
| Scale 1: | 6.5 (4.9) | 5.8 (5.1) | 5.4 (4.7) | 6.5 (4.3) | 8.3 (5.7) | 7.2 (5.3) |
| Somatization | 6.4 (5.0) | 5.6 (5.1) | 5.6 (4.9) | 6.4 (4.3) | 8.3 (5.6) | 7.1 (5.2) |
| Scale 2: | 11.9 (6.3) | 11.2 (6.0) | 11.0 (6.9) | 12.6 (5.8) | 12.3 (6.7) | 12.9 (5.7) |
| Depression | 11.9 (6.3) | 11.1 (5.9) | 11.1 (6.9) | 12.6 (5.9) | 12.5 (6.7) | 12.9 (5.6) |
| Scale 3: | 10.2 (5.7) | 9.0 (5.7) | 9.1 (6.0) | 10.4 (5.2) | 12.1 (6.2) | 10.9 (4.6) |
| Anxiety | 10.1 (5.8) | 8.9 (5.7) | 9.2 (6.1) | 10.3 (5.3) | 12.2 (6.2) | 10.9 (4.6) |

**Table 3:** Logistic regression with weighted deterioration in mental health as outcome (participants responding either 'Much worse' or 'Slightly worse' to the question: "How do you consider your mental health during the lockdown compared to the period before the corona pandemic came to Denmark?").

| *Clinical and demographic variables* | *OR* | *CI95%(OR)* | *z-value* | *P-value* |
|---|---|---|---|---|
| ***Sex (reference: male)*** | | | | |
| Female | 1.17 | (0.87;1.59) | 1.04 | 0.298 |
| ***Age (reference: ≤1st quantile of age)*** | | | | |
| 1st-2nd quantile | 1.02 | (0.66;1.58) | 0.11 | 0.912 |
| 2nd-3rd quantile | 0.61 | (0.37;1.01) | -1.90 | 0.057 |
| >3rd quantile | 0.66 | (0.37;1.17) | -1.43 | 0.153 |
| ***Civil status (reference: married & registered partnership)*** | | | | |
| Single | 0.99 | (0.62;1.58) | -0.05 | 0.963 |
| Divorced, Repeal of registered partnership | 1.00 | (0.60;1.67) | -0.01 | 0.992 |
| Widow/widower | 0.77 | (0.24;2.42) | -0.45 | 0.656 |
| ***Municipality (reference:* Inhabitants < 50,000** | | | | |
| Inhabitants 150,000-350,000*)* | 1.16 | (0.79;1.73) | 0.76 | 0.449 |
| Inhabitants 50,000-150,000 | 1.03 | (0.72;1.48) | 0.18 | 0.855 |
| ***Diagnosis (reference: F32-F33)*** | | | | |
| F10-F19 | 0.84 | (0.21;3.00) | -0.27 | 0.789 |
| F20-F29 | 0.57 | (0.37;0.87) | -2.58 | **0.010** |
| F30-F31 | 1.08 | (0.69;1.69) | 0.35 | 0.727 |
| F40-F49 | 1.46 | (0.93;2.30) | 1.62 | 0.105 |
| F50-F59 | 0.64 | (0.16;2.35) | -0.67 | 0.505 |
| F60-F69 | 1.41 | (0.73;2.78) | 1.01 | 0.311 |
| F90-F98 | 1.20 | (0.58;2.48) | 0.49 | 0.627 |
| Other F diagnoses | 0.72 | (0.17;2.70) | -0.48 | 0.629 |
| Z-diagnoses | 0.68 | (0.31;1.45) | -0.99 | 0.322 |
| ***Psychiatric admission pre-corona (reference: no admission)*** | | | | |
| *Admission* | 0.93 | (0.68;1.29) | -0.41 | 0.685 |
| ***Psychiatric outpatient visits pre-corona (reference: 1st quantile)*** | | | | |
| 1st-2nd quantile*, n (%)* | 1.24 | (0.83;1.85) | 1.04 | 0.300 |
| 2nd-3rd quantile*, n (%)* | 1.56 | (1.03;2.37) | 2.09 | **0.037** |
| >3rd quantile*, n (%)* | 1.45 | (0.92;2.29) | 1.59 | 0.112 |
| ***What is your highest attained educational level? (reference: long-cycle higher education)*** | | | | |
| Primary and lower secondary school | 1.15 | (0.64;2.07) | 0.48 | 0.634 |
| Upper secondary education | 1.31 | (0.75;2.28) | 0.95 | 0.344 |
| Skilled worker/craftsman | 1.07 | (0.53;2.14) | 0.19 | 0.852 |
| Short-cycle higher education | 1.13 | (0.64;2.00) | 0.41 | 0.681 |
| Medium-cycle higher education including bachelor | 1.20 | (0.69;2.08) | 0.64 | 0.521 |
| ***What describes your current employment status? (reference: full-time employed)*** | | | | |

| | | | | |
|---|---|---|---|---|
| Full-time student | 0.89 | (0.44;1.78) | -0.34 | 0.737 |
| Part-time student | 1.73 | (0.57;5.49) | 0.96 | 0.337 |
| Part-time employed | 0.87 | (0.35;2.09) | -0.32 | 0.748 |
| Subsidized employment | 1.95 | (0.94;4.08) | 1.78 | 0.076 |
| Absent owing to illness | 1.23 | (0.71;2.15) | 0.73 | 0.463 |
| Unemployed | 0.86 | (0.45;1.65) | -0.45 | 0.654 |
| Retired | 1.91 | (1.00;3.65) | 1.96 | 0.050 |
| ***Adults living in your household besides yourself? (reference: yes)*** | | | | |
| No | 1.39 | (1.01;1.91) | 2.04 | **0.042** |
| ***Children living in your household? (reference: yes)*** | | | | |
| No | 1.14 | (0.77;1.67) | 0.65 | 0.518 |
| ***Were you born in Denmark? (reference: yes)*** | | | | |
| No | 1.13 | (0.69;1.86) | 0.48 | 0.635 |

**Figure 3:** Perceived reasons for deterioration during the pandemic lockdown (weighted).

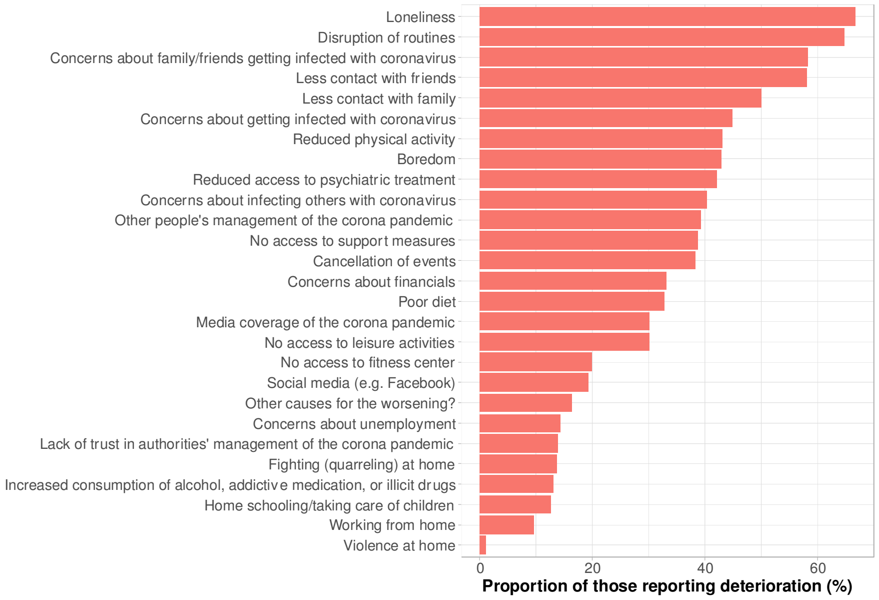

Perceived reasons for deterioration among the 492/960 (51%) of the respondents who reported that their overall mental health had worsened during the nationwide pandemic lockdown.

**Figure 4:** Perceived reasons for deterioration (weighted) among the 492/960 (51%) of the respondents who reported that their overall mental health had worsened during the nationwide pandemic lockdown stratified on the following diagnostic groups: psychotic disorder (ICD-10: F2x), bipolar disorder (ICD-10: F30-31), unipolar depression (ICD-10: F32-33), anxiety disorder (ICD-10: F4x), and personality disorder (ICD-10: F6x). The following categories are removed due to microdata in the subdiagnosis groups: “Home schooling / taking care of children”, “Working from home”, Fighting (quarreling) at home”, “Violence at home”, “Increased consumption of alcohol, addictive medication, or illicit drugs”, “Concerns about unemployment”, “Other causes for the worsening?” have been removed from the figure due to small numbers and risk of identification of individuals.

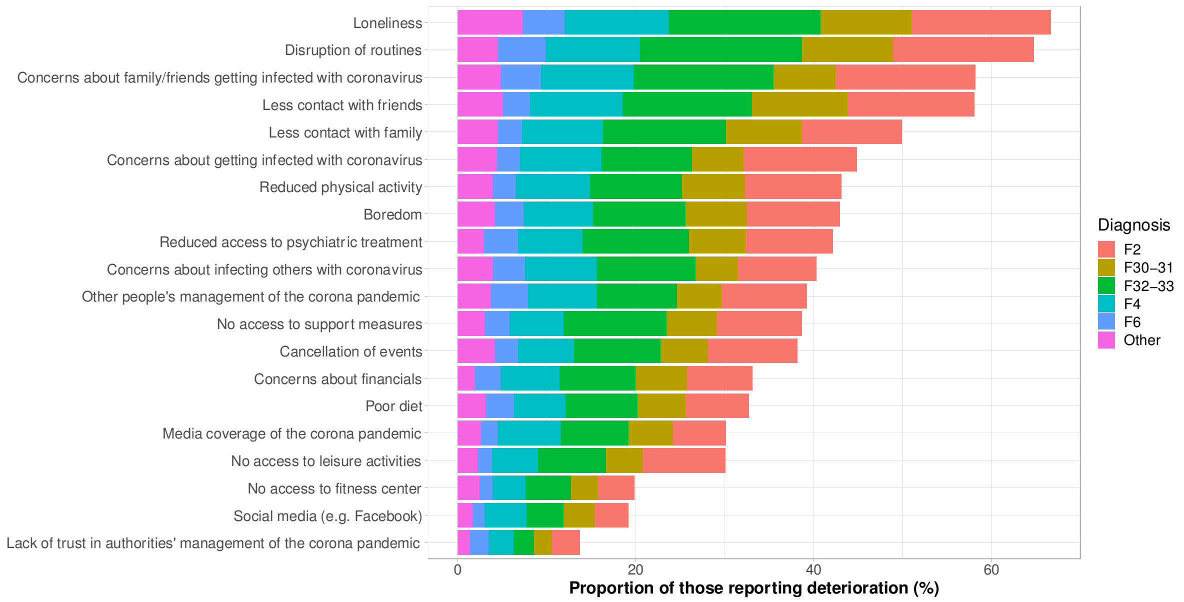

**Table 4:** Perceived reasons for deterioration (weighted) among the 492/960 (51%) respondents who reported that their overall mental health had worsened during the nationwide pandemic lockdown and stratified on the following diagnostic groups: psychotic disorder (ICD-10: F2x), bipolar disorder (ICD-10: F30-31), unipolar depression (ICD-10: F32-33), anxiety disorder (ICD-10: F4x), and personality disorder (ICD-10: F6x). "-" indicates microdata.

| ***Deterioration in mental health*** *%* | ***Respondents*** *(n = 492)* | ***F2*** | ***F30-F31*** | ***F32-F33*** | ***F4*** | ***F6*** | ***Other*** |
|---|---|---|---|---|---|---|---|
| Concerns about getting infected with coronavirus | 44.9 | 12.8 | 5.8 | 10.2 | 9.2 | 2.6 | 4.4 |
| Concerns about family/friends getting infected with coronavirus | 58.3 | 15.8 | 7.0 | 15.7 | 10.4 | 4.5 | 4.8 |
| Concerns about infecting others with coronavirus | 40.3 | 8.8 | 4.8 | 11.1 | 8.0 | 3.7 | 3.9 |
| Lack of trust in authorities' management of the corona pandemic | 13.8 | 3.2 | 2.0 | 2.2 | 2.8 | 2.1 | 1.4 |
| Home schooling/taking care of children | 12.6 | - | - | - | - | - | - |
| Working from home | 9.6 | - | - | - | - | - | - |
| No access to support measures | 38.7 | 9.6 | 5.6 | 11.6 | 6.0 | 2.8 | 3.0 |
| No access to leisure activities | 30.1 | 9.3 | 4.1 | 7.6 | 5.2 | 1.5 | 2.3 |
| No access to fitness center | 19.9 | 4.1 | 3.0 | 5.1 | 3.8 | 1.4 | 2.5 |
| Reduced physical activity | 43.1 | 10.7 | 7.1 | 10.3 | 8.5 | 2.6 | 3.9 |
| Reduced access to psychiatric treatment | 42.2 | 9.8 | 6.4 | 11.9 | 7.3 | 3.8 | 3.0 |

| | | | | | | | |
|---|---|---|---|---|---|---|---|
| Disruption of routines | 64.8 | 15.9 | 10.2 | 18.2 | 10.6 | 5.3 | 4.5 |
| Cancellation of events | 38.2 | 10.1 | 5.4 | 9.7 | 6.3 | 2.6 | 4.2 |
| Media coverage of the corona pandemic | 30.1 | 6.0 | 4.9 | 7.6 | 7.1 | 1.9 | 2.6 |
| Social media (e.g. Facebook) | 19.2 | 3.8 | 3.5 | 4.2 | 4.6 | 1.4 | 1.7 |
| Other people's management of the corona pandemic | 39.3 | 9.7 | 5.1 | 9.0 | 7.7 | 4.2 | 3.7 |
| Fighting (quarreling) at home | 13.7 | - | - | - | - | - | - |
| Violence at home | 1.1 | - | - | - | - | - | - |
| Increased consumption of alcohol, addictive medication, or illicit drugs | 13.0 | - | - | - | - | - | - |
| Concerns about unemployment | 14.3 | - | - | - | - | - | - |
| Concerns about financials | 33.2 | 7.4 | 5.8 | 8.5 | 6.6 | 2.9 | 1.9 |
| Boredom | 43.0 | 10.5 | 6.9 | 10.4 | 7.8 | 3.2 | 4.2 |
| Less contact with friends | 58.1 | 14.3 | 10.7 | 14.6 | 10.4 | 3.1 | 5.0 |
| Less contact with family | 49.9 | 11.2 | 8.6 | 13.8 | 9.2 | 2.7 | 4.5 |
| Loneliness | 66.7 | 15.7 | 10.2 | 17.1 | 11.7 | 4.8 | 7.3 |
| Poor diet | 32.8 | 7.2 | 5.4 | 8.1 | 5.7 | 3.3 | 3.1 |
| Other causes for the worsening? | 16.3 | - | - | - | - | - | - |

**Figure 5:** Weighted response to the 14 questions focusing on changes in mental health and compliance to treatment during the COVID-19 lockdown of Denmark in the spring of 2020.

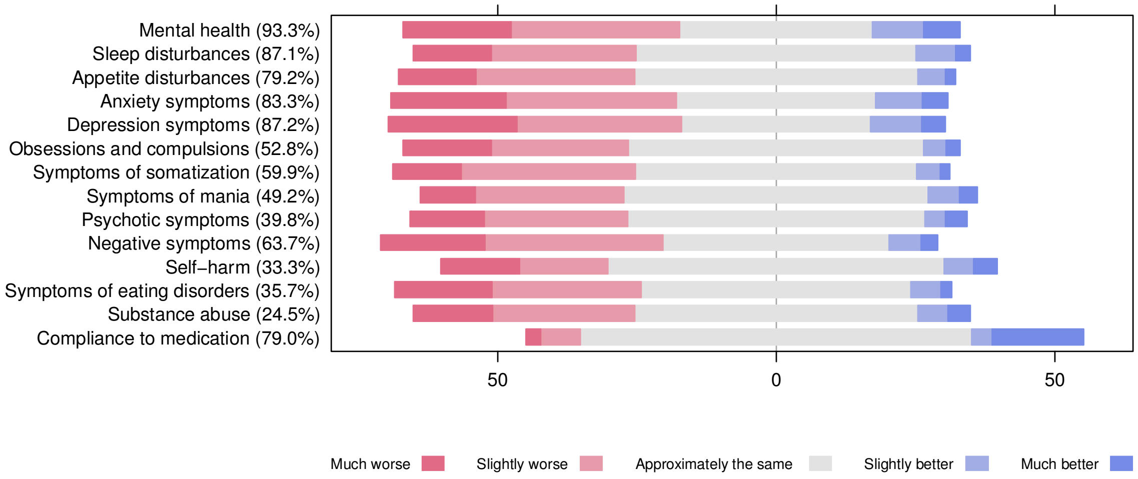

The numbers in the parentheses represent the proportion of respondents endorsing one of the five likert-scale responses (they could also choose: "The symptom was not present (either before or during the lockdown)" or choose not to respond).

### Figure 6: Psychotic disorders

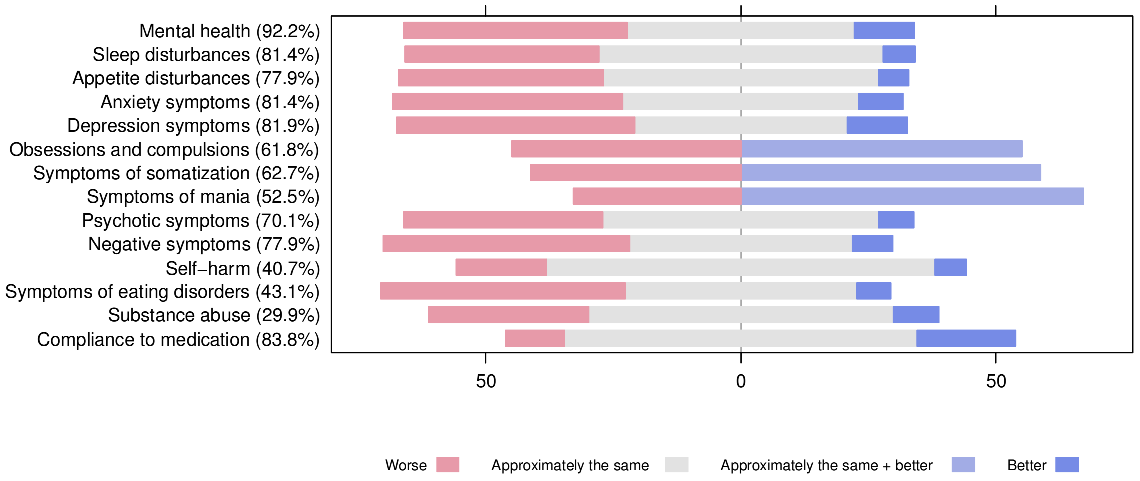

### Figure 7: Bipolar disorder

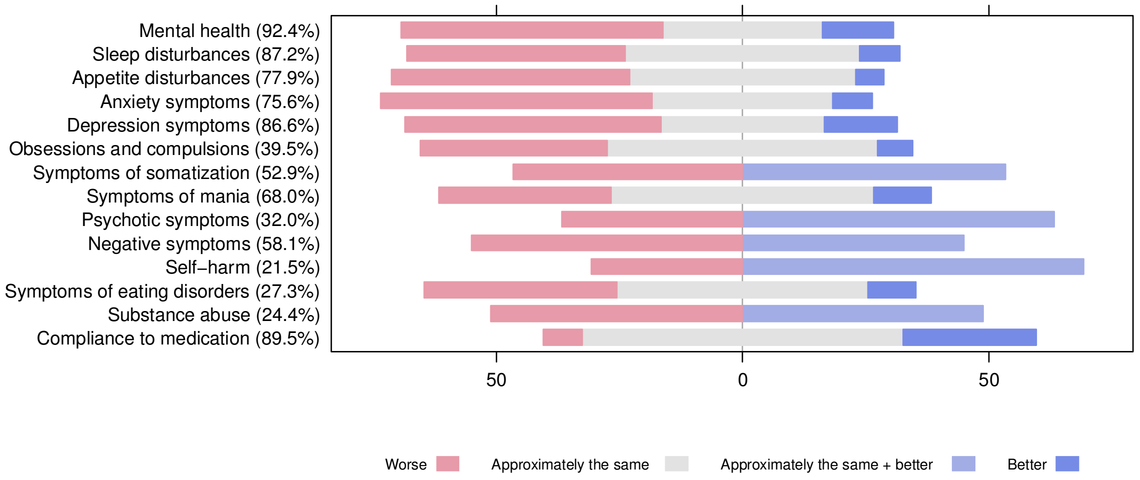

### Figure 8: Unipolar depression

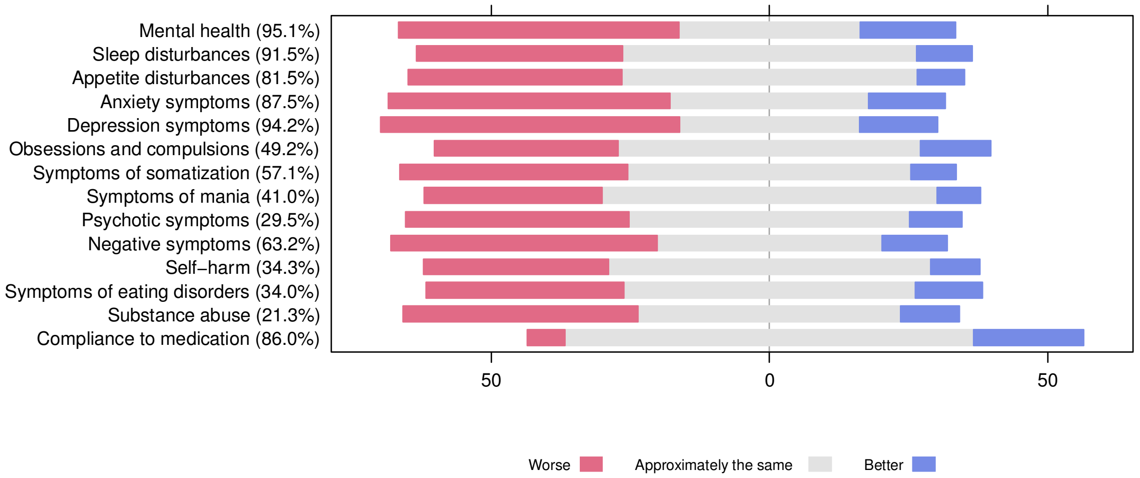

**Figure 9: Anxiety- and stress-related disorders**

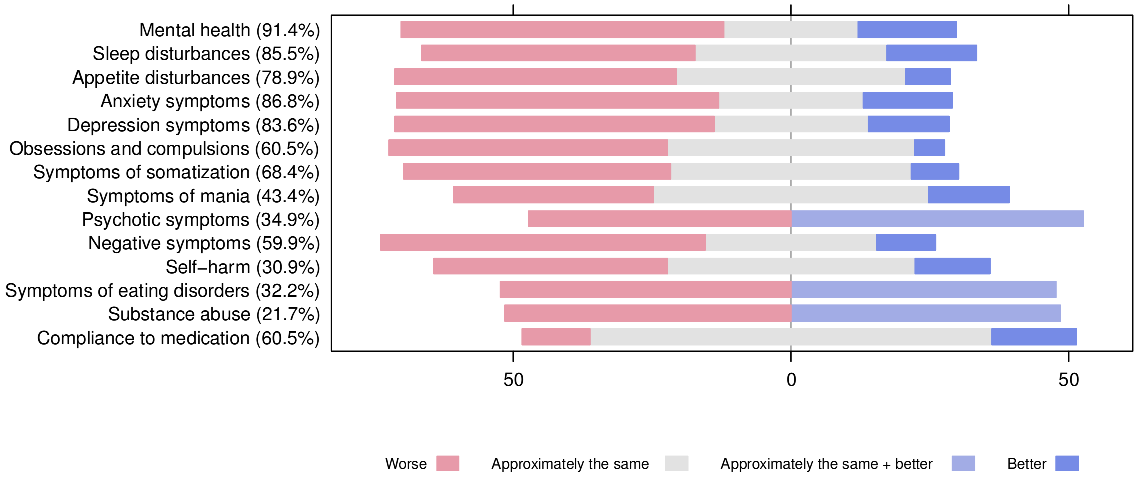

**Figure 10: Personality disorders**

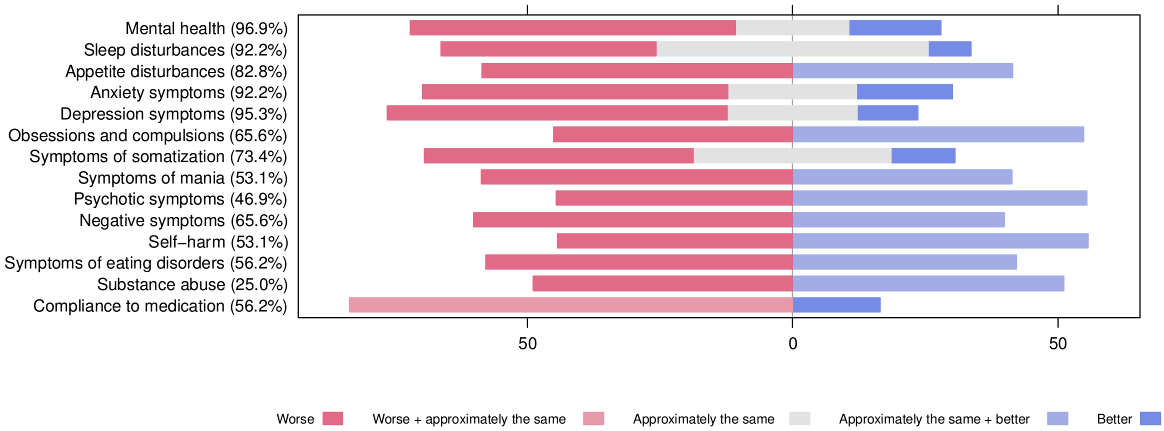

**Table 5:** Proportions of response to the 14 mental health questions for all the respondents and stratified on the following diagnostic groups: psychotic disorder (ICD-10: F2x), bipolar disorder (ICD-10: F30-31), unipolar depression (ICD-10: F32-33), anxiety disorder (ICD-10: F4x), and personality disorder (ICD-10: F6x). "-" indicates microdata.

| *All respondents (n=1029)* % | *Much worse* | *Slightly worse* | *Approximately the same* | *Slightly better* | *Much better* |
|---|---|---|---|---|---|
| Mental health (93.2%) | 19.5 | 30.2 | 34.6 | 9.2 | 6.5 |
| Sleep disturbances (86.9%) | 14.0 | 26.0 | 50.2 | 7.1 | 2.7 |
| Appetite disturbances (79.1%) | 14.0 | 28.4 | 50.8 | 5.0 | 1.8 |
| Anxiety symptoms (83.3%) | 20.7 | 30.6 | 35.8 | 8.3 | 4.6 |
| Depression symptoms (87.1%) | 23.0 | 29.6 | 33.9 | 9.2 | 4.2 |
| Obsessions and compulsions (52.5%) | 15.9 | 24.6 | 53.0 | 4.0 | 2.5 |
| Symptoms of somatization (59.8%) | 12.3 | 31.3 | 50.5 | 4.2 | 1.7 |
| Symptoms of mania (49.4%) | 9.9 | 26.7 | 54.6 | 5.6 | 3.2 |
| Psychotic symptoms (40.2%) | 13.4 | 25.7 | 53.3 | 3.7 | 4.0 |
| Negative symptoms (63.9%) | 18.8 | 31.9 | 40.6 | 5.7 | 3.0 |
| Self-harm (33.4%) | 14.2 | 15.8 | 60.4 | 5.2 | 4.3 |
| Symptoms of eating disorders (36.0%) | 17.4 | 26.8 | 48.4 | 5.4 | 2.0 |
| Substance abuse (24.7%) | 14.2 | 25.5 | 50.8 | 5.5 | 4.0 |
| Compliance to medication (78.8%) | 2.6 | 7.1 | 70.3 | 3.6 | 16.5 |

| ***F2 (n=204)***<br>*%* | ***Worse*** | ***Approximately the same*** | ***Approximately the same + better*** | ***Better*** |
|---|---|---|---|---|
| Mental health (92.2%) | 43.8 | 44.6 | - | 11.7 |
| Sleep disturbances (81.4%) | 38.0 | 55.7 | - | 6.3 |
| Appetite disturbances (77.9%) | 40.1 | 53.9 | - | 5.9 |
| Anxiety symptoms (81.4%) | 45.0 | 46.3 | - | 8.7 |
| Depression symptoms (81.9%) | 46.6 | 41.6 | - | 11.9 |
| Obsessions and compulsions (61.8%) | 44.9 | - | 55.1 | - |
| Symptoms of somatization (62.7%) | 41.3 | - | 58.7 | - |
| Symptoms of mania (52.5%) | 32.8 | - | 67.2 | - |
| Psychotic symptoms (70.1%) | 39.1 | 54.0 | - | 6.9 |
| Negative symptoms (77.9%) | 48.3 | 43.7 | - | 7.9 |
| Self-harm (40.7%) | 17.7 | 76.2 | - | 6.1 |
| Symptoms of eating disorders (43.1%) | 47.9 | 45.4 | - | 6.7 |
| Substance abuse (29.9%) | 31.3 | 59.7 | - | 8.9 |
| Compliance to medication (83.8%) | 11.5 | 69.2 | - | 19.3 |

| ***F30-31 (n=172)***<br>*%* | ***Worse*** | ***Approximately the same*** | ***Approximately the same + better*** | ***Better*** |
|---|---|---|---|---|
| Mental health (92.4%) | 53.1 | 32.4 | - | 14.5 |
| Sleep disturbances (87.2%) | 44.3 | 47.6 | - | 8.1 |
| Appetite disturbances (77.9%) | 48.3 | 45.9 | - | 5.7 |
| Anxiety symptoms (75.6%) | 55.2 | 36.6 | - | 8.1 |
| Depression symptoms (86.6%) | 52.0 | 33.1 | - | 14.9 |
| Obsessions and compulsions (39.5%) | 38.0 | 54.9 | - | 7.1 |
| Symptoms of somatization (52.9%) | 46.6 | - | 53.4 | - |
| Symptoms of mania (68.0%) | 35.0 | 53.3 | - | 11.7 |
| Psychotic symptoms (32.0%) | 36.7 | - | 63.3 | - |
| Negative symptoms (58.1%) | 55.0 | - | 45.0 | - |
| Self-harm (21.5%) | 30.7 | - | 69.3 | - |
| Symptoms of eating disorders (27.3%) | 39.2 | 51.0 | - | 9.7 |
| Substance abuse (24.4%) | 51.1 | - | 48.9 | - |
| Compliance to medication (89.5%) | 7.8 | 65.2 | - | 27.1 |

| ***F32-33 (n=329)***<br>*%* | ***Worse*** | ***Approximately the same*** | ***Better*** |
|---|---|---|---|
| Mental health (95.1%) | 50.4 | 32.6 | 17.1 |
| Sleep disturbances (91.5%) | 37.1 | 52.8 | 10.0 |
| Appetite disturbances (81.5%) | 38.4 | 53.0 | 8.6 |
| Anxiety symptoms (87.5%) | 50.6 | 35.7 | 13.7 |
| Depression symptoms (94.2%) | 53.7 | 32.4 | 14.0 |
| Obsessions and compulsions (49.2%) | 33.1 | 54.3 | 12.6 |
| Symptoms of somatization (57.1%) | 41.0 | 50.9 | 8.1 |
| Symptoms of mania (41.0%) | 32.0 | 60.1 | 7.9 |
| Psychotic symptoms (29.5%) | 40.2 | 50.4 | 9.4 |
| Negative symptoms (63.2%) | 47.9 | 40.3 | 11.8 |
| Self-harm (34.3%) | 33.3 | 57.8 | 8.9 |
| Symptoms of eating disorders (34.0%) | 35.6 | 52.3 | 12.1 |
| Substance abuse (21.3%) | 42.3 | 47.2 | 10.5 |
| Compliance to medication (86.0%) | 6.8 | 73.4 | 19.8 |

| ***F4 (n=152)***<br>*%* | ***Worse*** | ***Approximately the same*** | ***Approximately the same + better*** | ***Better*** |
|---|---|---|---|---|
| Mental health (91.4%) | 58.2 | 24.1 | - | 17.7 |
| Sleep disturbances (85.5%) | 49.3 | 34.5 | - | 16.2 |
| Appetite disturbances (78.9%) | 50.7 | 41.2 | - | 8.1 |
| Anxiety symptoms (86.8%) | 58.0 | 26.0 | - | 16.0 |
| Depression symptoms (83.6%) | 57.5 | 27.8 | - | 14.6 |

| | | | | | |
|---|---|---|---|---|---|
| Obsessions and compulsions (60.5%) | 50.1 | 44.5 | - | 5.4 | |
| Symptoms of somatization (68.4%) | 48.0 | 43.4 | - | 8.5 | |
| Symptoms of mania (43.4%) | 36.0 | 49.5 | - | 14.5 | |
| Psychotic symptoms (34.9%) | 47.3 | - | 52.7 | - | |
| Negative symptoms (59.9%) | 58.5 | 30.8 | - | 10.7 | |
| Self-harm (30.9%) | 42.0 | 44.6 | - | 13.5 | |
| Symptoms of eating disorders (32.2%) | 52.3 | - | 47.7 | - | |
| Substance abuse (21.7%) | 51.5 | - | 48.5 | - | |
| Compliance to medication (60.5%) | 12.3 | 72.3 | - | 15.3 | |
| ***F6 (n=64)*** *%* | ***Worse*** | ***Worse + approximately the same*** | ***Approximately the same*** | ***Approximately the same + better*** | ***Better*** |
| Mental health (96.9%) | 61.3 | - | 21.6 | - | 17.1 |
| Sleep disturbances (92.2%) | 40.6 | - | 51.5 | - | 7.9 |
| Appetite disturbances (82.8%) | 58.6 | - | - | 41.4 | - |
| Anxiety symptoms (92.2%) | 57.5 | - | 24.6 | - | 17.8 |
| Depression symptoms (95.3%) | 64.1 | - | 24.7 | - | 11.2 |
| Obsessions and compulsions (65.6%) | 45.1 | - | - | 54.9 | - |
| Symptoms of somatization (73.4%) | 50.7 | - | 37.5 | - | 11.8 |
| Symptoms of mania (53.1%) | 58.7 | - | - | 41.3 | - |
| Psychotic symptoms (46.9%) | 44.6 | - | - | 55.4 | - |
| Negative symptoms (65.6%) | 60.1 | - | - | 39.9 | - |
| Self-harm (53.1%) | 44.3 | - | - | 55.7 | - |
| Symptoms of eating disorders (56.2%) | 57.8 | - | - | 42.2 | - |
| Substance abuse (25.0%) | 48.9 | - | - | 51.1 | - |
| Compliance to medication (56.2%) | - | 83.5 | - | - | 16.5 |
