## Supplement 2 for "Mental health of patients with mental illness during the COVID-19 pandemic lockdown: A questionnaire-based survey weighted for attrition"

**Supplementary Figure 1:** Visualization of the non-weighted and weighted mean and 95%-confidence interval of the WHO-5 total score, the BSI-18 total score, the three subscales of BSI-18.

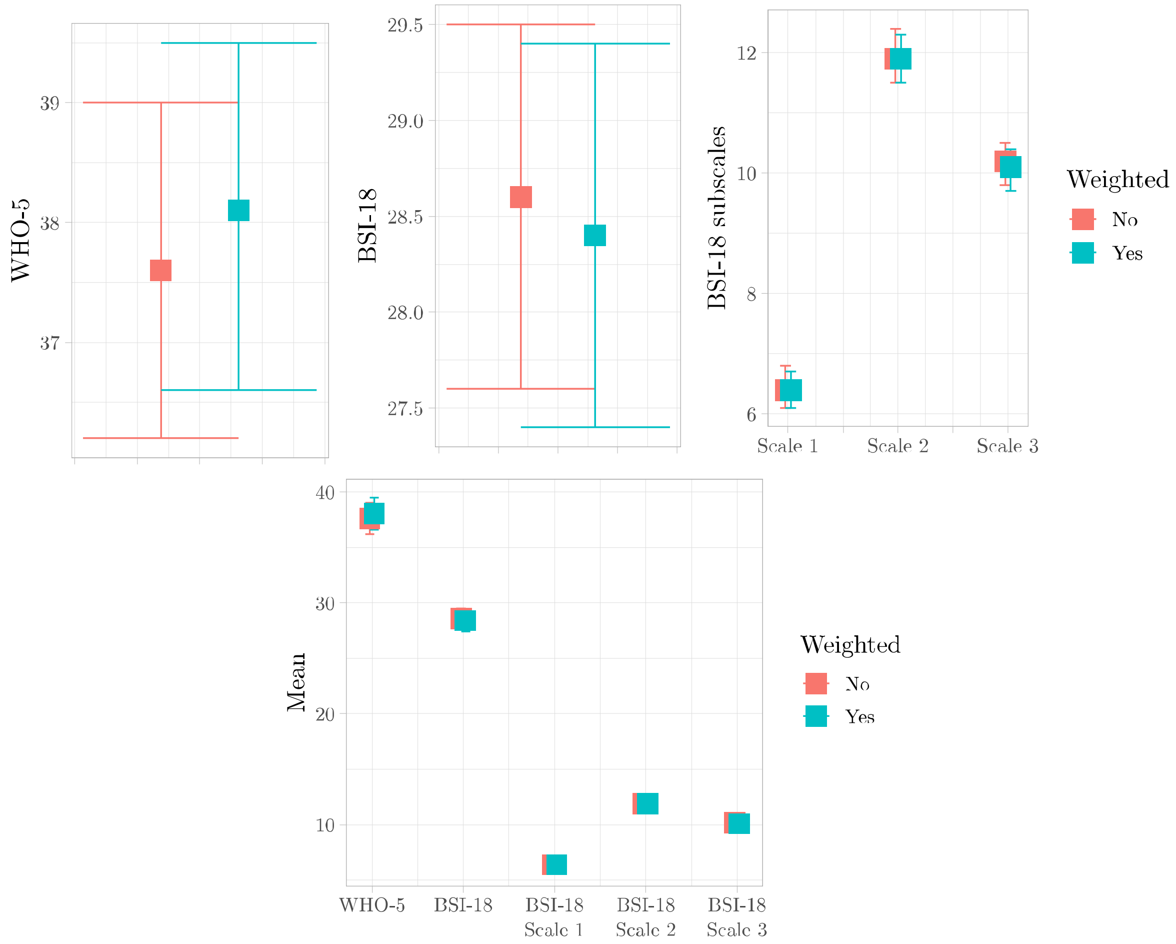


**Supplementary Table 1:** Table displaying the non-weighted (red) and weighted (blue) mean (SD) of the WHO-5 total score, the BSI-18 total score, the three subscales of BSI-18 for all respondents and stratified on the ICD-10 diagnosis: F2, F30-F31, F32-F33, F4 and F6.

| ***Questionnaires***<br>*Mean (SD)* | ***Respondents***<br>*(n = 992)* | ***F2***<br>*(n = 204)* | ***F30-F31***<br>*(n = 172)* | ***F32-F33***<br>*(n = 329)* | ***F4***<br>*(n = 152)* | ***F6***<br>*(n = 64)* |
|---|---|---|---|---|---|---|
| ***WHO-5*** | | | | | | |
| Total score* | 37.6 (22.5)<br>38.1 (22.7) | 42.9 (23.3)<br>42.9 (23.1) | 38.6 (23.5)<br>38.4 (23.6) | 34.1 (20.8)<br>34.1 (20.9) | 35.8 (22.1)<br>35.1 (22.1) | 34.9 (19.8)<br>34.9 (20.0) |
| ***BSI-18*** | | | | | | |
| Total score | 28.6 (15.0)<br>28.4 (15.2) | 25.9 (14.9)<br>25.6 (14.8) | 25.5 (15.5)<br>25.9 (15.9) | 29.5 (13.4)<br>29.4 (13.5) | 32.7 (16.9)<br>33.1 (16.7) | 31.0 (13.6)<br>31.0 (13.4) |
| Scale 1:<br>Somatization | 6.4 (4.9)<br>6.4 (5.0) | 5.8 (5.1)<br>5.6 (5.1) | 5.4 (4.7)<br>5.6 (4.9) | 6.5 (4.3)<br>6.4 (4.3) | 8.3 (5.7)<br>8.3 (5.6) | 7.2 (5.3)<br>7.1 (5.2) |
| Scale 2:<br>Depression | 11.9 (6.3)<br>11.9 (6.3) | 11.2 (6.0)<br>11.1 (5.9) | 11.0 (6.9)<br>11.1 (6.9) | 12.6 (5.8)<br>12.6 (5.9) | 12.3 (6.7)<br>12.6 (6.7) | 12.9 (5.7)<br>12.9 (5.6) |
| Scale 3:<br>Anxiety | 10.2 (5.7)<br>10.1 (5.8) | 9.0 (5.7)<br>8.9 (5.7) | 9.1 (6.0)<br>9.2 (6.1) | 10.4 (5.2)<br>10.4 (5.3) | 12.1 (6.2)<br>12.2 (6.2) | 10.9 (4.6)<br>10.9 (4.6) |

**Supplementary Figure 2:** Perceived reasons for deterioration (weighted) among the 479/925 (52%) of the respondents who reported that their overall mental health had worsened during the nationwide pandemic lockdown stratified on the following diagnostic groups: psychotic disorder (ICD-10: F2x), bipolar disorder (ICD-10: F30-31), unipolar depression (ICD-10: F32-33), anxiety disorder (ICD-10: F4x), and personality disorder (ICD-10: F6x). The following categories are removed due to microdata in the subdiagnosis groups: "Home schooling / taking care of children", "Working from home", Fighting (quarreling) at home", "Violence at home", "Increased consumption of alcohol, addictive medication, or illicit drugs", "Concerns about unemployment", "Other causes for the worsening?" have been removed from the figure due to small numbers and risk of identification of individuals.

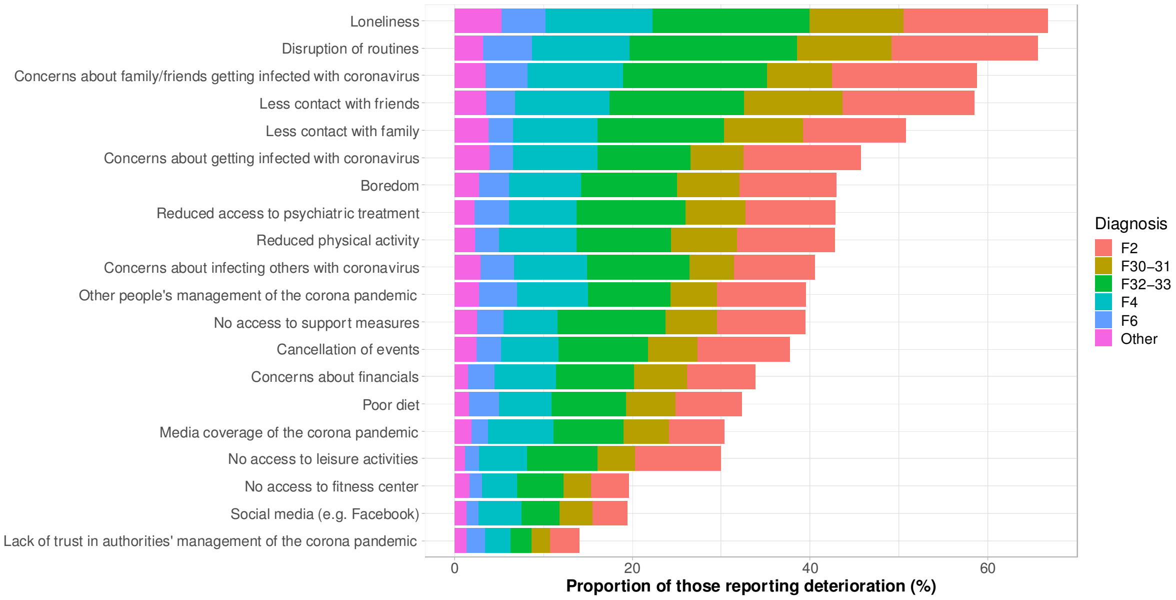


**Supplementary Table 2:** Perceived reasons for deterioration (weighted) among the 479/925 (52%) respondents who reported that their overall mental health had worsened during the nationwide pandemic lockdown and stratified on the following diagnostic groups: psychotic disorder (ICD-10: F2x), bipolar disorder (ICD-10: F30-31), unipolar depression (ICD-10: F32-33), anxiety disorder (ICD-10: F4x), and personality disorder (ICD-10: F6x).

| *Deterioration in mental health %* | *Respondents (n = 479)* | *F2* | *F30-F31* | *F32-F33* | *F4* | *F6* | *Other* |
|---|---|---|---|---|---|---|---|
| Concerns about getting infected with coronavirus | 45.7 | 13.2 | 6.0 | 10.5 | 9.5 | 2.7 | 3.9 |
| Concerns about family/friends getting infected with coronavirus | 58.8 | 16.3 | 7.3 | 16.2 | 10.7 | 4.7 | 3.5 |
| Concerns about infecting others with coronavirus | 40.6 | 9.2 | 5.0 | 11.5 | 8.2 | 3.8 | 2.9 |
| Lack of trust in authorities' management of the corona pandemic | 14.1 | 3.4 | 2.1 | 2.3 | 2.9 | 2.1 | 1.3 |
| Home schooling/taking care of children | 12.4 | - | - | - | - | - | - |
| Working from home | 9.7 | - | - | - | - | - | - |

| | | | | | | | |
|---|---|---|---|---|---|---|---|
| No access to support measures | 39.5 | 9.9 | 5.8 | 12.1 | 6.2 | 2.9 | 2.5 |
| No access to leisure activities | 30.0 | 9.6 | 4.3 | 7.9 | 5.4 | 1.6 | 1.1 |
| No access to fitness center | 19.6 | 4.2 | 3.1 | 5.2 | 3.9 | 1.4 | 1.7 |
| Reduced physical activity | 42.8 | 11.1 | 7.4 | 10.7 | 8.7 | 2.7 | 2.3 |
| Reduced access to psychiatric treatment | 42.9 | 10.1 | 6.7 | 12.4 | 7.5 | 3.9 | 2.2 |
| Disruption of routines | 65.6 | 16.5 | 10.6 | 18.9 | 11.0 | 5.5 | 3.2 |
| Cancellation of events | 37.7 | 10.4 | 5.6 | 10.1 | 6.5 | 2.7 | 2.5 |
| Media coverage of the corona pandemic | 30.3 | 6.2 | 5.1 | 7.9 | 7.3 | 1.9 | 1.9 |
| Social media (e.g. Facebook) | 19.5 | 4.0 | 3.7 | 4.3 | 4.8 | 1.4 | 1.3 |
| Other people's management of the corona pandemic | 39.6 | 10.0 | 5.2 | 9.3 | 8.0 | 4.3 | 2.7 |
| Fighting (quarreling) at home | 14.2 | - | - | - | - | - | - |
| Violence at home | 1.1 | - | - | - | - | - | - |
| Increased consumption of alcohol, addictive medication, or illicit drugs | 13.2 | - | - | - | - | - | - |
| Concerns about unemployment | 14.4 | - | - | - | - | - | - |
| Concerns about financials | 33.8 | 7.6 | 6.0 | 8.8 | 6.9 | 3.0 | 1.5 |
| Boredom | 42.9 | 10.9 | 7.1 | 10.8 | 8.1 | 3.3 | 2.8 |
| Less contact with friends | 58.5 | 14.8 | 11.1 | 15.1 | 10.7 | 3.2 | 3.5 |
| Less contact with family | 50.8 | 11.6 | 8.9 | 14.2 | 9.5 | 2.7 | 3.8 |
| Loneliness | 66.7 | 16.2 | 10.6 | 17.7 | 12.0 | 4.9 | 5.3 |
| Poor diet | 32.3 | 7.4 | 5.6 | 8.4 | 5.9 | 3.4 | 1.6 |
| Other causes for the worsening? | 16.7 | - | - | - | - | - | - |

### Supplementary Figure 3: Psychotic disorders

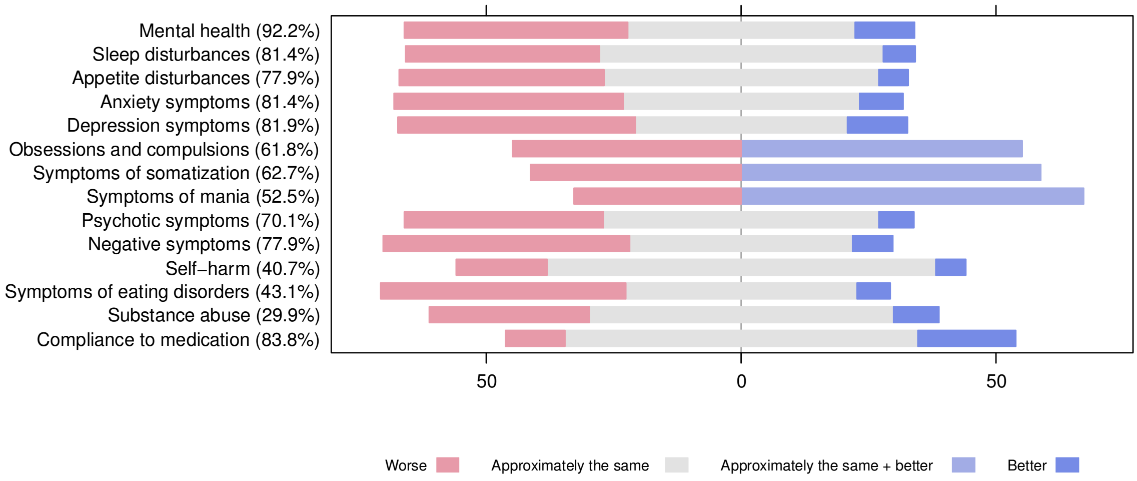


### Supplementary Figure 4: Bipolar disorder

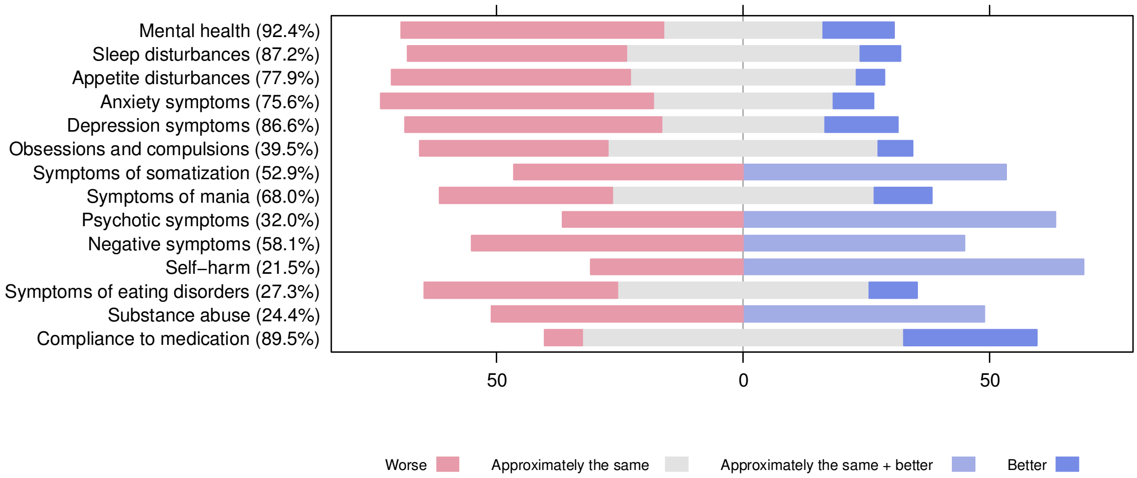


### Supplementary Figure 5: Unipolar depression

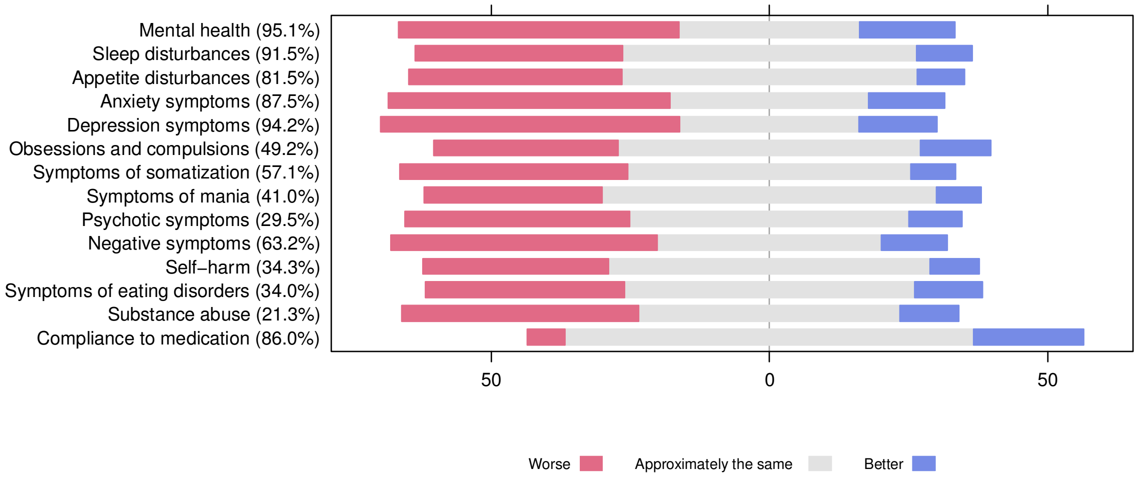

**Supplementary Figure 6: Anxiety- and stress-related disorders**

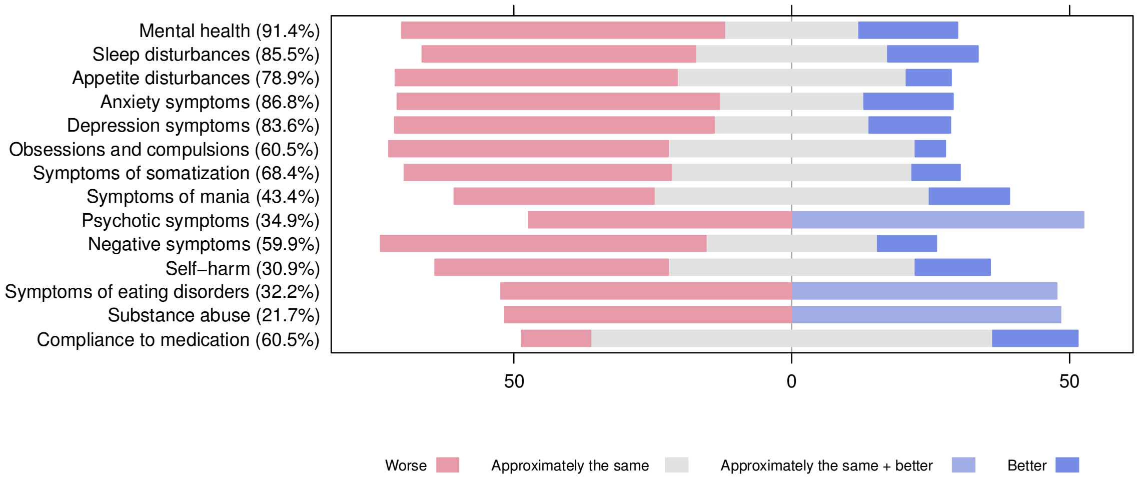


**Supplementary Figure 7: Personality disorders**

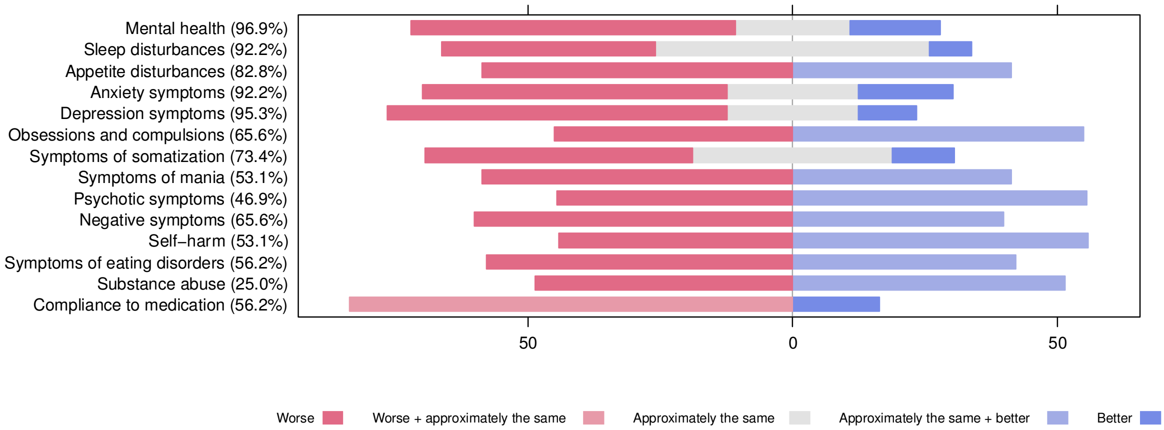


**Supplementary Table 3:** Proportions of response to the 14 mental health questions for all the respondents and stratified on the following diagnostic groups: psychotic disorder (ICD-10: F2x), bipolar disorder (ICD-10: F30-31), unipolar depression (ICD-10: F32-33), anxiety disorder (ICD-10: F4x), and personality disorder (ICD-10: F6x).

| *All respondents (n=992)* % | *Much worse* | *Slightly worse* | *Approximately the same* | *Slightly better* | *Much better* |
|---|---|---|---|---|---|
| Mental health (93.2%) | 19.6 | 30.8 | 34.4 | 8.9 | 6.3 |
| Sleep disturbances (86.9%) | 14.2 | 26.4 | 49.6 | 7.0 | 2.9 |
| Appetite disturbances (79.1%) | 14.4 | 29.0 | 50.1 | 5.2 | 1.4 |
| Anxiety symptoms (83.3%) | 20.8 | 30.3 | 36.3 | 8.2 | 4.4 |
| Depression symptoms (87.1%) | 23.0 | 29.9 | 34.0 | 9.0 | 4.0 |
| Obsessions and compulsions (52.5%) | 16.5 | 24.9 | 52.0 | 3.9 | 2.7 |
| Symptoms of somatization (59.8%) | 12.4 | 31.4 | 50.0 | 4.3 | 1.8 |
| Symptoms of mania (49.4%) | 10.1 | 27.0 | 54.5 | 5.0 | 3.4 |
| Psychotic symptoms (40.2%) | 13.9 | 26.1 | 52.3 | 3.6 | 4.1 |
| Negative symptoms (63.9%) | 19.2 | 32.3 | 39.9 | 5.5 | 3.1 |
| Self-harm (33.4%) | 14.6 | 16.1 | 60.9 | 4.4 | 4.1 |
| Symptoms of eating disorders (36.0%) | 17.2 | 27.2 | 48.4 | 5.2 | 2.1 |
| Substance abuse (24.7%) | 14.6 | 25.5 | 50.5 | 5.2 | 4.2 |
| Compliance to medication (78.8%) | 2.7 | 7.2 | 70.1 | 3.6 | 16.4 |

| ***F2 (n=204)***<br>*%* | ***Worse*** | ***Approximately the same*** | ***Approximately the same + better*** | ***Better*** |
|---|---|---|---|---|
| Mental health (92.2%) | 43.8 | 44.6 | - | 11.7 |
| Sleep disturbances (81.4%) | 38.1 | 55.6 | - | 6.3 |
| Appetite disturbances (77.9%) | 40.2 | 53.8 | - | 5.9 |
| Anxiety symptoms (81.4%) | 45.0 | 46.3 | - | 8.6 |
| Depression symptoms (81.9%) | 46.6 | 41.6 | - | 11.8 |
| Obsessions and compulsions (61.8%) | 44.9 | - | 55.1 | - |
| Symptoms of somatization (62.7%) | 41.3 | - | 58.7 | - |
| Symptoms of mania (52.5%) | 32.8 | - | 67.2 | - |
| Psychotic symptoms (70.1%) | 39.1 | 54.0 | - | 6.9 |
| Negative symptoms (77.9%) | 48.4 | 43.7 | - | 7.9 |
| Self-harm (40.7%) | 17.8 | 76.2 | - | 6.0 |
| Symptoms of eating disorders (43.1%) | 48.1 | 45.3 | - | 6.6 |
| Substance abuse (29.9%) | 31.4 | 59.7 | - | 8.9 |
| Compliance to medication (83.8%) | 11.6 | 69.2 | - | 19.2 |

| ***F30-31 (n=172)***<br>*%* | ***Worse*** | ***Approximately the same*** | ***Approximately the same + better*** | ***Better*** |
|---|---|---|---|---|
| Mental health (92.4%) | 53.2 | 32.3 | - | 14.5 |
| Sleep disturbances (87.2%) | 44.4 | 47.5 | - | 8.1 |
| Appetite disturbances (77.9%) | 48.4 | 45.9 | - | 5.7 |
| Anxiety symptoms (75.6%) | 55.3 | 36.6 | - | 8.1 |
| Depression symptoms (86.6%) | 52.1 | 33.1 | - | 14.9 |
| Obsessions and compulsions (39.5%) | 38.2 | 54.8 | - | 7.0 |
| Symptoms of somatization (52.9%) | 46.6 | - | 53.4 | - |
| Symptoms of mania (68.0%) | 35.1 | 53.2 | - | 11.7 |
| Psychotic symptoms (32.0%) | 36.7 | - | 63.3 | - |
| Negative symptoms (58.1%) | 55.1 | - | 44.9 | - |
| Self-harm (21.5%) | 30.9 | - | 69.1 | - |
| Symptoms of eating disorders (27.3%) | 39.2 | 51.0 | - | 9.8 |
| Substance abuse (24.4%) | 51.1 | - | 48.9 | - |
| Compliance to medication (89.5%) | 7.8 | 65.2 | - | 27.0 |

| ***F32-33 (n=329)***<br>*%* | ***Worse*** | ***Approximately the same*** | ***Better*** |
|---|---|---|---|
| Mental health (95.1%) | 50.4 | 32.5 | 17.1 |
| Sleep disturbances (91.5%) | 37.2 | 52.8 | 10.0 |
| Appetite disturbances (81.5%) | 38.3 | 53.0 | 8.6 |
| Anxiety symptoms (87.5%) | 50.7 | 35.6 | 13.7 |
| Depression symptoms (94.2%) | 53.7 | 32.3 | 14.0 |
| Obsessions and compulsions (49.2%) | 33.2 | 54.3 | 12.6 |
| Symptoms of somatization (57.1%) | 41.0 | 50.8 | 8.1 |
| Symptoms of mania (41.0%) | 32.0 | 60.0 | 8.0 |
| Psychotic symptoms (29.5%) | 40.4 | 50.2 | 9.5 |
| Negative symptoms (63.2%) | 47.9 | 40.2 | 11.9 |
| Self-harm (34.3%) | 33.4 | 57.7 | 8.9 |
| Symptoms of eating disorders (34.0%) | 35.7 | 52.1 | 12.2 |
| Substance abuse (21.3%) | 42.5 | 47.0 | 10.5 |
| Compliance to medication (86.0%) | 6.8 | 73.4 | 19.8 |

| ***F4 (n=152)***<br>*%* | ***Worse*** | ***Approximately the same*** | ***Approximately same + better*** | ***Better*** |
|---|---|---|---|---|

| | | | | | |
|---|---|---|---|---|---|
| Mental health (91.4%) | 58.1 | 24.2 | - | 17.7 | |
| Sleep disturbances (85.5%) | 49.2 | 34.6 | - | 16.2 | |
| Appetite disturbances (78.9%) | 50.7 | 41.2 | - | 8.1 | |
| Anxiety symptoms (86.8%) | 57.9 | 26.1 | - | 16.0 | |
| Depression symptoms (83.6%) | 57.5 | 27.9 | - | 14.6 | |
| Obsessions and compulsions (60.5%) | 50.2 | 44.5 | - | 5.4 | |
| Symptoms of somatization (68.4%) | 48.0 | 43.4 | - | 8.6 | |
| Symptoms of mania (43.4%) | 36.0 | 49.5 | - | 14.5 | |
| Psychotic symptoms (34.9%) | 47.4 | - | 52.6 | - | |
| Negative symptoms (59.9%) | 58.6 | 30.8 | - | 10.7 | |
| Self-harm (30.9%) | 42.0 | 44.5 | - | 13.5 | |
| Symptoms of eating disorders (32.2%) | 52.3 | - | 47.7 | - | |
| Substance abuse (21.7%) | 51.6 | - | 48.4 | - | |
| Compliance to medication (60.5%) | 12.4 | 72.4 | - | 15.3 | |
| ***F6 (n=64)*** *%* | ***Worse*** | ***Worse + approximately the same*** | ***Approximately the same*** | ***Approximately the same + better*** | ***Better*** |
| Mental health (96.9%) | 61.3 | - | 21.6 | - | 17.1 |
| Sleep disturbances (92.2%) | 40.4 | - | 51.7 | - | 7.9 |
| Appetite disturbances (82.8%) | 58.7 | - | - | 41.3 | - |
| Anxiety symptoms (92.2%) | 57.5 | - | 24.7 | - | 17.9 |
| Depression symptoms (95.3%) | 64.2 | - | 24.7 | - | 11.1 |
| Obsessions and compulsions (65.6%) | 45.0 | - | - | 55.0 | - |
| Symptoms of somatization (73.4%) | 50.6 | - | 37.7 | - | 11.7 |
| Symptoms of mania (53.1%) | 58.7 | - | - | 41.3 | - |
| Psychotic symptoms (46.9%) | 44.5 | - | - | 55.5 | - |
| Negative symptoms (65.6%) | 60.1 | - | - | 39.9 | - |
| Self-harm (53.1%) | 44.2 | - | - | 55.8 | - |
| Symptoms of eating disorders (56.2%) | 57.8 | - | - | 42.2 | - |
| Substance abuse (25.0%) | 48.6 | - | - | 51.4 | - |
| Compliance to medication (56.2%) | - | 83.6 | - | - | 16.4 |

### Overview of missing data

**Response-rate for the questions that are not conditioned upon each other:**
**"Adults living in your household besides yourself", "Children living in your household", "Were you born in Denmark?", "What is your highest attained educational level?", "What describes your current employment status?", "WHO-5", "BSI-18", "14 mental health questions" (21 categories)**

7% have answered 100% of the questionnaire
13% have answered 90% of the questionnaire (removed mental health 11,13)
23% have answered 81% of the questionnaire (removed mental health 9,11:13)
34% have answered 71% of the questionnaire (removed mental health 6,8:9,11:13)
59% have answered 62% of the questionnaire (removed mental health 6:13)
76% have answered 52% of the questionnaire (removed mental health 3,6:13)
86% have answered 43% of the questionnaire (removed mental health 2:4,6:13)
93% have answered 33% of the questionnaire (removed mental health 1:13)
97% have answered 24% of the questionnaire (removed mental health 1:13, WHO-5 and BSI-18)

**Response-rate to socio-demographics:** 97%

- Adults living in your household besides yourself (99%)
- Children living in your household (99%)
- Were you born in Denmark? (98%)
- What is your highest attained educational level? (98%)
- What describes your current employment status? (98%)

**Response-rate to WHO-5 and BSI-18:** 94%

- WHO-5: 97%
- BSI-18: 95%

**Response-rate to mental health:**

- Deterioration: 52%
- The same: 32%
- Improvement: 16%

**Response-rate to the 14 mental health questions:** 8%

- Mental health (93.2%)
- Sleep disturbances (86.9%)
- Appetite disturbances (79.1%)
- Anxiety symptoms (83.3%)
- Depression symptoms (87.1%)
- Obsessions and compulsions (52.5%)
- Symptoms of somatization (59.8%)
- Symptoms of mania (49.4%)
- Psychotic symptoms (40.2%)
- Negative symptoms (63.9%)
- Self-harm (33.4%)
- Symptoms of eating disorders (36.0%)
- Substance abuse (24.7%)
- Compliance to medication (78.8%)
