## Supplement 1 for "Mental health of patients with mental illness during the COVID-19 pandemic lockdown: A questionnaire-based survey weighted for attrition"

| COVID-19 Survey Questionnaire (English version) | | |
|---|---|---|
| | **Background.**<br>**We begin with some questions about your background.** | **Options separated by commas** |
| 1 | How many adults are living in your household, besides yourself? | 0, 1, 2, 3, 4, 5, or more |
| 2 | How many children are living in your household? | 0, 1, 2, 3, 4, 5, or more |
| 3 | Were you born in Denmark? | Yes, No |
| 4 | (Conditional, if question 3 = No)<br>Where were you born? | Africa, North America, South America, Asia, Middle East, Oceania (Australia, New Zealand, etc.), Europe |
| 5 | What is your highest attained educational level? | Primary and lower secondary school,<br>Higher general, preparatory, or commercial examination programmes (upper secondary education),<br>Skilled worker/craftsman,<br>Short-cycle higher education (less than 3 years of study),<br>Medium-cycle higher education (3-4 years of study),<br>Long-cycle higher education (more than 4 years of study) |
| 6 | What describes your current employment status the best? | Student (full time), Student (part time), Full-time employed, Part-time employed, Subsidized employment, Absent owing to illness, Unemployed, Retired |
| 6 | **The 5-item World Health Organization Well-Being Index (WHO-5)**[a] | |
| 7 | **The 18-item Brief Symptom Inventory (BSI-18)**[b] | |
| 8 | **Mental health during the corona pandemic.**<br>**The last questions are about your mental health during the corona pandemic.**<br><br>**From March 11 to April 14, 2020, the corona pandemic peaked in Denmark. Schools and institutions were closed, public employees who did not carry out critical functions were sent home, all non-essential travels were advised against, and the Danish borders were closed. We now ask you to think back on that period, which we will refer to as "the lockdown".**<br><br>**In the following questions, we would like you to consider whether the symptoms (signs of illness you may or may not have experienced DURING THE LOCKDOWN worsened, improved, or were approximately at the same level compared to the period BEFORE THE CORONA PANDEMIC came to Denmark.**<br><br>**Example 1: If you experienced that the symptom in question remained the same during the lockdown compared to the period before the corona pandemic came to Denmark, reply "Approximately the same level"**<br><br>**Example 2: If you have not previously experienced the symptom in question, but you have experienced it during the lockdown, reply either "Increased/worsened to a lesser degree" or "Increased/worsened to a greater degree" – depending on the severity of the symptom.** | |

| | | |
|---|---|---|
| 9 | **Mental health.**<br>How would you assess your mental health during the lockdown compared to the period before the corona pandemic came to Denmark? | Much worse,<br>Slightly worse,<br>Approximately the same,<br>Slightly better,<br>Much better |
| | **Sleep disturbances.**<br>Sleep disturbances may include decreased or increased sleep duration as well as poor sleep quality. This could, for instance, be difficulties falling asleep, waking up during nighttime, or waking up early in the morning. It may also be an increased need for sleep and daytime sleepiness, not explained by a preceding lack of sleep. | |
| 10 | How has the severity of your supposed sleep disturbances been during the lockdown compared to the period before the corona pandemic came to Denmark? | Increased/worsened to a greater degree,<br>Increased/worsened to a lesser degree,<br>Approximately the same level,<br>Decreased/improved to a small degree,<br>Decreased/improved to a large degree,<br>The symptom was not present (either before or during the lockdown) |
| | **Appetite disturbances.**<br>Appetite disturbances may be both increased or decreased appetite accompanied by weight gain or weight loss, respectively. | |
| 11 | How has the severity of your supposed appetite disturbance been during the lockdown compared to the period before the corona pandemic came to Denmark? | Same as 10 |
| | **Anxiety symptoms.**<br>Anxiety symptoms include a general feeling of being anxious, worried, tense, or feeling keyed up inside. It may also come in attacks of high intensity and shorter duration, or it can be associated with thoughts about specific things or situations. This may be accompanied by trembling, sweating, heart palpitations, dizziness, shortness of breath, or a feeling of tightness in the throat. | |
| 12 | How has the severity of your supposed anxiety symptoms been during the lockdown compared to the period before the corona pandemic came to Denmark? | Same as 10 |
| | **Depression symptoms.**<br>Depression symptoms include a feeling of sadness, guilt, decreased pleasure or interest in things and/or people, lack of energy, increased tiredness, reduced self-esteem, self-blame, thoughts of death, trouble concentrating, sleep disturbances, and weight changes. | |
| 13 | How has the severity of your supposed depression symptoms been during the lockdown compared to the period before the corona pandemic came to Denmark? | Same as 10 |
| | **Obsessions and compulsions.**<br>Obsessions are repeated/intrusive, unpleasant thoughts or inner images, often concerning contamination, illness, sex, or violence. Compulsions are repetitive behaviors or rituals one feels driven to perform, often not intuitively coherent, e.g. an urge to wash hands, count things, control doors, locks, or cooking plates, or carry out actions in a certain way. | |

| | | |
|---|---|---|
| 14 | How has the severity of your supposed obsessions or compulsions been during the lockdown compared to the period before the corona pandemic came to Denmark? | Same as 10 |
| | **Symptoms of somatization.**<br>Somatization is characterized by a preoccupation with changing physical symptoms accompanied by concerns about serious affection of one's body (e.g. severe disease). Examples of physical symptoms include pain (e.g. joint, muscle, or abdominal pain), nausea, bloating, vomiting, diarrhea, shortness of breath, chest pains, pricking, tingling, and numb sensations or sensory disturbances. | |
| 15 | How has the severity of your supposed symptoms of somatization been during the lockdown compared to the period before the corona pandemic came to Denmark? | Same as 10 |
| | **Symptoms of mania.**<br>Mania is a condition with elevated mood (e.g. feeling excited, angry, or irritated) that may be accompanied by restlessness, physical unrest, decreased need for sleep, or increased sex drive. Disinhibition, grandiose thoughts, unusual talkativeness, or increased energy leading to overactivity may also occur. | |
| 16 | How has the severity of your supposed symptoms of mania been during the lockdown compared to the period before the corona pandemic came to Denmark? | Same as 10 |
| | **Psychotic symptoms.**<br>Psychosis is characterized by a disturbed sense of reality. This may include a sense of being watched or talked about in a certain way by others, and that they are out to get you, feeling persecuted or under surveillance, possessing exceptional abilities that others do not possess, being chosen for a special role by higher powers, sensing that others can access your thoughts, or having different convictions that are not shared by others. Psychosis can also present itself as sensory experiences, like hearing voices or sounds whilst being alone, or seeing, tasting, or smelling something that others cannot see, taste, or smell (hallucinations). | |
| 17 | How has the severity of your supposed symptoms of psychosis been during the lockdown compared to the period before the corona pandemic came to Denmark? | Same as 10 |
| | **Negative symptoms.**<br>In psychotic disorders, like schizophrenia, so-called negative symptoms often occur. Negative symptoms cover social isolation/withdrawal, lack of initiative, apathy, fewer, and diminished mood swings (e.g. difficulties feeling pleasure or sorrow), decreased interest in activities/hobbies, and reduced attention to personal care. | |
| 18 | How has the severity of your supposed negative symptoms been during the lockdown compared to the period before the corona pandemic came to Denmark? | Same as 10 |
| | **Self-harm.**<br>Self-harm comprises hitting, cutting, biting, or burning oneself, or other actions that cause physical damage to oneself. The actions may be an attempt to relieve inner tension or make unpleasant thoughts or emotions disappear. Self-harm may also be present in | |

| | | |
|---|---|---|
| | a compulsory form in which the same action is repeated (e.g. pulling out hair or scratching oneself). | |
| 19 | How has the severity of your supposed self-harm been during the lockdown compared to the period before the corona pandemic came to Denmark? | Same as 10 |
| | **Symptoms of eating disorders (e.g. anorexia, bulimia, or binge-eating disorder).**<br>Symptoms of eating disorders include fear of gaining weight or an exaggerated focus on losing weight. This may be accompanied by excessive physical activity, use of laxatives, diuretics, self-induced forced vomiting, or restricted food consumption. Loss of control, leading to excess eating, may also occur. | |
| 20 | How has the severity of your supposed symptoms of eating disturbance been during the lockdown compared to the period before the corona pandemic came to Denmark? | Same as 10 |
| | **Substance abuse.**<br>Substance abuse is defined as use of substances that influences one to such a degree (physically, mentally, or socially) that one's daily activities are affected. Alcohol, hash, benzodiazepines, whippits (nitrous oxide/"laughing gas"), amphetamine, cocaine, or the like, are all examples of substances that may be abused. Signs of substance abuse could be considering cutting down on the use, irritability towards others commenting on one's substance use, feeling guilty for consuming the substance, or needing the substance in order to get through the day, relax, or sleep at night. | |
| 21 | How has the severity of your supposed substance abuse been during the lockdown compared to the period before the corona pandemic came to Denmark? | Same as 10 |
| | **Prescribed medication for mental disorders.**<br>The last question is about the medication prescribed by your doctor as part of the treatment for your mental disorder (if any). We wish to examine whether you have taken the medication as prescribed by your doctor (right dosage and frequency). There may be many reasons for not taking one's medication as prescribed by the doctor, e.g. forgetting it, experiencing adverse effects, or maybe considering the medication to be unnecessary. In your answer, we ask you to ignore the reason why you have not taken the medication as prescribed by your doctor (if so), and only consider to what extent you have followed the instructions provided by your doctor. | |
| 22 | To what extent have you followed your doctors' instructions regarding the intake of medication for mental disorder (dosage and frequency) during the lockdown compared to the period before the corona pandemic came to Denmark? | To a much lesser degree,<br>To a slightly lesser degree,<br>To approximately the same degree,<br>To a slightly greater degree,<br>To a much greater degree,<br>I do not receive medication for a mental disorder |
| 23 | (Conditional, if question 31 = "Much worse" OR "Slightly worse")<br><br>**Worsening of mental health.**<br>In the question regarding your mental health during the lockdown, you replied that it had worsened. What causes do you | • Concerns about getting infected with coronavirus,<br>• Concerns about family/friends getting infected with coronavirus,<br>• Concerns about infecting others with coronavirus, |

| | | |
|---|---|---|
| | attribute the worsening in your mental health during the lockdown to (mark all the causes you find relevant)? | • Lack of trust in the authorities' handling of the corona pandemic,<br>• Home schooling / taking care of children,<br>• Working from home,<br>• No access to support measures,<br>• No access to leisure activities,<br>• No access to fitness center,<br>• Reduced physical activity,<br>• Poorer access to psychiatric treatment,<br>• Disruption of routines,<br>• Cancellation of events,<br>• Media coverage of the corona pandemic,<br>• Social media (e.g. Facebook),<br>• Other people's handling of the corona pandemic,<br>• Fighting (quarreling) at home,<br>• Violence at home,<br>• Increased consumption of alcohol, addictive medication, or illicit drugs,<br>• Concerns about unemployment,<br>• Concerns about finances,<br>• Boredom,<br>• Less contact with friends,<br>• Less contact with family,<br>• Loneliness,<br>• Poor diet,<br>• Other causes for the worsening? |
| 24 | (Conditional, if question 45 = "Other causes for the worsening") Please state the other causes you attribute to the worsening of your mental health. | [Text input] |
| 25 | (Conditional, if question 31 = "Much better" OR "Slightly better")<br><br>**Improvement of mental health.**<br>In the question regarding your mental health during the lockdown, you replied that it had improved. What causes do you attribute the improvement in your mental health during the lockdown to (mark all the causes you find relevant)? | • Fewer demands from the local and public authorities (job center, municipality, etc.),<br>• Fewer demands from employer,<br>• Fewer demands from family/friends,<br>• Cancellation of events,<br>• Less social activity,<br>• Less noise,<br>• Less stigmatization,<br>• Working from home,<br>• Home schooling / taking care of children,<br>• More leisure time,<br>• Better access to psychiatric treatment,<br>• Better diet,<br>• Social media (e.g. Facebook),<br>• Increased exercise/workout, |

| | | |
|---|---|---|
| | | • Experiences in the nature,<br>• Fewer arguments/less fighting at home,<br>• Reduced consumption of alcohol, addictive medication, or illicit drugs,<br>• Other causes for the improvement? |
| 26 | (Conditional, if question 47 = “Other causes for the improvement?”)<br>Please state the other causes you attribute to the worsening of your mental health. | [Text input] |

[a] Topp CW, Østergaard SD, Søndergaard S, Bech P. The WHO-5 Well-Being Index: a systematic review of the literature. Psychotherapy and psychosomatics. 2015;84(3):167-176.
[b] Zabora J, BrintzenhofeSzoc K, Jacobsen P, Curbow B, Piantadosi S, Hooker C, Owens A, Derogatis L. A new psychosocial screening instrument for use with cancer patients. Psychosomatics. 2001;42(3):241-6.
